# Molecular and Proteomic Profiles of Radioiodine Refractory Papillary Thyroid Cancer

**DOI:** 10.1101/2024.09.22.24314143

**Authors:** Hanqing Liu, Jiaxi Wang, Yan Zhou, Pingping Hu, Lu Li, Dan Yang, Deguang Kong, Zhiliang Xu, Yaoting Sun, Chuang Chen

**Affiliations:** Department of Breast and Thyroid Surgery, Renmin Hospital of Wuhan University, Wuhan University at Jiefang Road 238, Wuhan 430060, China; School of Medicine, Westlake University, Hangzhou 310024, Zhejiang Province, China; Westlake Center for Intelligent Proteomics, Westlake Laboratory of Life Sciences and Biomedicine, Hangzhou 310024, Zhejiang Province, China; Research Center for Industries of the Future, School of Life Sciences, Westlake University, Hangzhou 310024, Zhejiang Province, China; College of Pharmaceutical Sciences, Zhejiang University, Hangzhou 310024, China; Department of Cardiology, the Second Affiliated Hospital of Chongqing Medical University, Chongqing 400010, China

**Author notes:** Equal contribution (Hanqing Liu, Jiaxi Wang, Yan Zhou). Correspondence: Prof. Chuang Chen, M.D., Department of Thyroid and Breast Surgery, Renmin Hospital of Wuhan University, Wuhan University at Jiefang Road 238, Wuhan 430060, RP China. Dr. Yaoting Sun, Ph.D., Westlake Laboratory of Life Sciences and Biomedicine, School of Medicine, Westlake University (Yunqi Campus) at No. 18 Shilongshan Rd., Hangzhou 310024, PR China.

**Keywords:** thyroid carcinoma, radioactive iodine, refractoriness, proteomics, NGS, *RET/PTC3*

## Abstract

**Background:** Despite the generally favorable prognosis of papillary thyroid cancers (PTCs) following surgery with/without radioactive iodine (RAI) therapy, approximately one-third of patients experiencing recurrence and metastasis eventually develop resistance to RAI, leading to poor outcomes. However, the mechanisms underlying RAI-refractoriness remain elusive. This study aimed to assess the molecular and proteomic characteristics of RAI-refractory PTC (RR-PTC) for deeper insights.

**Methods:** The medical records were reviewed for the selection and grouping of RR-PTC patients and RAI-sensitive controls. RR-PTC patients were divided into three subgroups: continuous RAI uptake (ID), loss of uptake at the first I-131 treatment (iDF) and lost gradually (iDG). Proteomic profiling and targeted next-generation sequencing were performed on primary lesions. The incidence of gene mutations and fusions was compared across groups. Bioinformatic analysis was subsequently conducted to identify the differentially expressed proteins and enriched pathways.

**Results:** Forty-eight PTC patients with recurrence and/or metastasis were included. The expression profiles of the RR-PTC and control groups were similar. In the subgroup comparison, enriched pathways related to MAPK and TNF signaling were associated with negative I-131 uptake and tumor tolerance with positive I-131 uptake. The *BRAF*^V600E^ mutation was less common in the ID group, whereas the *TERT* promoter mutation was more common in the iDF group. *NCOA4*-*RET* fusion was more common in the ID group.

**Conclusion:** The present study constructed the first proteomic profile of RR-PTC. The identified proteins and pathways may be promising biomarkers and drug targets. Gene alterations can aid in the early diagnosis of RR-PTC.

## 1 Introduction

Thyroid cancer ranks eighth among female malignancies (1). Its incidence has increased threefold or greater during the past four decades in many countries (2, 3). Papillary thyroid cancer (PTC) is the most common histological type and accounts for 70-96% of all thyroid cancers (3, 4). Most patients with PTC achieve complete remission after undergoing lobectomy or total thyroidectomy with/without neck dissection. However, approximately half of PTC patients present with aggressive tumor behaviors, such as extrathyroidal extension, aggressive histology, vascular invasion, multiple lymph node metastases, and distant metastases (5, 6). These patients have an intermediate or high risk of recurrence and are thus considered or recommended for postsurgical radioactive iodine (RAI) therapy (7). Unfortunately, in case of recurrence or metastasis, one to two-thirds of patients in this subset ultimately develop RAI-refractoriness (RAIR), especially those with a high risk of recurrence (8). RAI therapy barely improves patient prognosis under these conditions. The 10-year survival rate of patients with RAI-refractory PTC (RR-PTC) has decreased to 20-65%, and their life expectancy has substantially decreased (8, 9).

Currently, the diagnosis of RR-PTC depends on an I-131 whole-body scan (WBS) and evidence of regional and/or distant metastatic lesions (7). Owing to the high affinity of thyroid follicular cells for iodine, radioactive isotopes are concentrated in PTC cells and exert a tumor-killing effect by emitting β-rays. In most cases, two or more I-131 WBSs are required for the final confirmation of RAIR. Given that the routine time interval between two I-131 WBSs is six months (10), the diagnosis of RAIR is made nine to twelve months after the initial surgery. The time lag postpones the start of systemic therapy (e.g. multi-kinase inhibitors, MKIs) and/or local treatments. Early identification of the RAIR can avoid unnecessary RAI therapy and thus improve patient prognosis by shifting to alternative treatments.

Many studies have focused on the early diagnosis of RAIR cancer during the past decade. Positron emission tomography (PET) is a valuable tool. Previous studies have shown that the negative I-131 and positive F-18-FDG uptake are indicative of the RAIR (11). In addition to radiological tools, *BRAF*^V600E^ and *TERT* promoter mutations can also contribute to the diagnosis of RAIR. A previous retrospective study revealed that a combination of *BRAF*^V600E^ and *TERTp* mutations, which presented in one-fifth of the PTC cohort, predicts RR-PTC at a positive predictive value of 97.4% (12). However, for approximately 80% of patients without the genetic duet, the negative predictive value was only 47.6%. A subsequent study revealed that 52.9% of patients with the genetic duet presented with RAIR (13). Generally, several factors, including high cost and unsatisfactory accuracy, have restricted the use of these diagnostic tools in clinical practice. In addition, although the RAIR is related to gene alterations in clinical studies (14, 15), the mechanism underlying refractoriness is not fully understood. For example, the NIS protein was not necessarily downregulated. It can be upregulated and exert a non-pump pro-tumorigenic effect on thyroid cancer cells (16).

Proteins are important participants in tumorigenic processes and dedifferentiation. Mass-based proteomics is a rapidly developing technique that facilitates comprehensive investigations into molecular activities and biological processes involved in cancer initiation and progression. Recently, proteomics has been used in several areas of thyroid cancer research, including discrimination of benign and malignant nodules, subtype identification, and risk stratification (17, 18). During the past two years, Sun *et al.* constructed a comprehensive thyroid tissue proteomic spectral library and distinguished between follicular adenomas and follicular thyroid carcinomas (19). In addition, Shi *et al.* plotted the multi-omic atlas of medullary thyroid carcinoma via the proteomic technique (20). The proteomic profile of RR-PTC has not yet been revealed, which may help improve the early diagnosis of RR-PTC in clinical practice and understand the mechanism of RAIR.

This study aimed to construct 23-gene panel-based molecular and proteomic profiles of RR-PTC and to identify promising biomarkers related to RAIR, which could aid in early identification and drug target selection.

### 2 Results

#### 2.1 Patient cohort

Forty-eight patients with locoregional or distant metastatic PTCs were included in our study (Table 1). The patients in the RR-PTC group were slightly older than those in the control group (*P* = 0.421). Interestingly, all five patients ≥ 55 years old were in the RR-PTC group. Although females were more common in the control group (72.7% *vs.* 54.1%, *P* = 0.319), the difference was not significant. In addition, no differences were found in the tumor diagnosis, smoking status or alcohol consumption, body mass index or the incidence of concomitant thyroid diseases between the two groups.

**Table 1.**
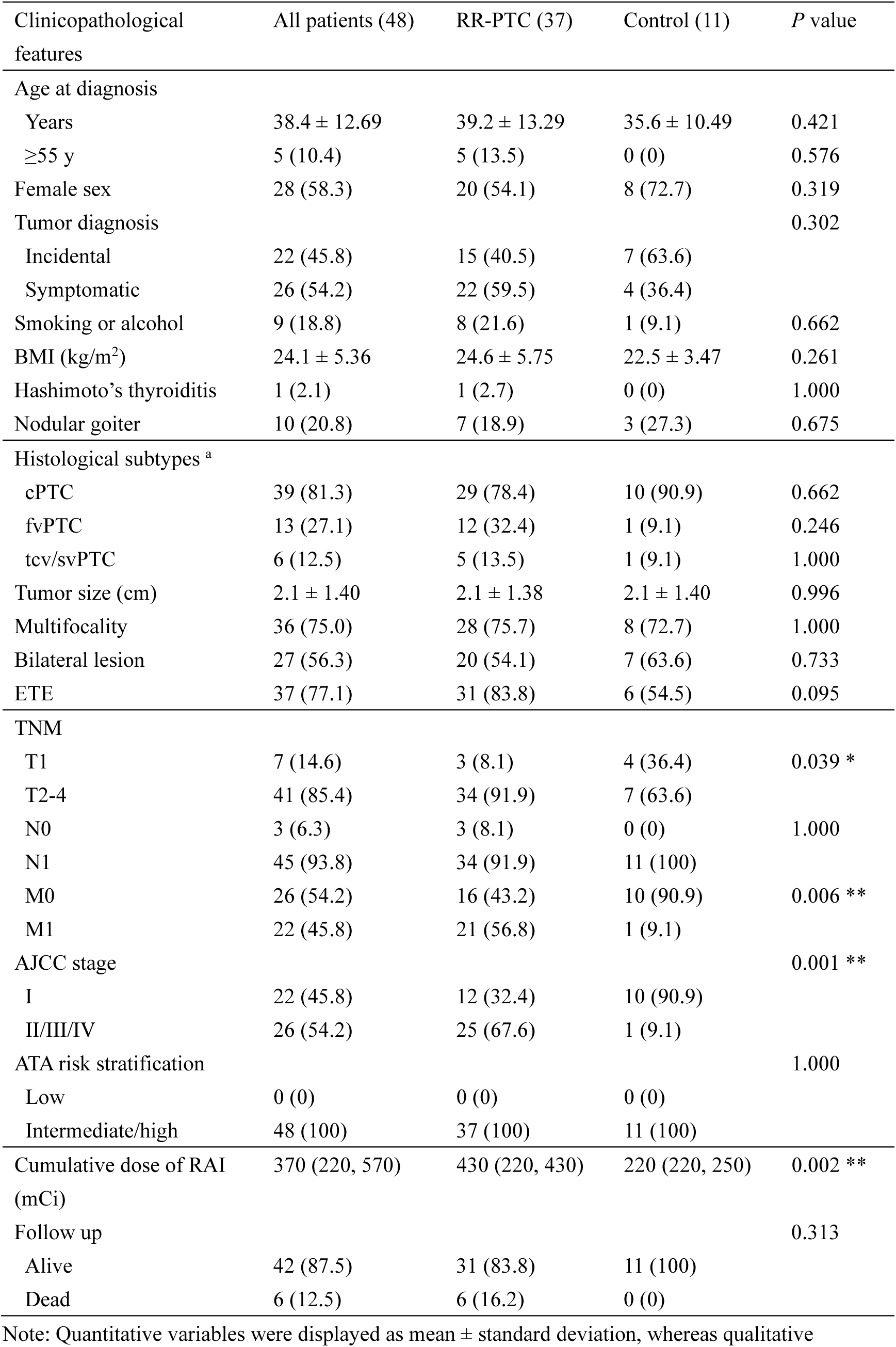
Demographic and clinicopathologic characteristic of included patients

The histopathological subtypes of PTC were identically distributed between the two groups. No significant differences were found in terms of tumor size, multifocality or bilateral lesions. The incidence of extrathyroidal extension (ETE) was greater in the RR-PTC group with marginal statistical significance (*P* = 0.095). Owing to differences in age and ETE, T2-4 categories were more common in patients with RR-PTC (91.9% *vs.* 63.6%, *P* = 0.039). The presence of lymph node metastatic lesion (LNM) was similar in the two groups, but distant metastasis was much more common in the patients with RR-PTC (56.8% *vs.* 9.1%, *P* = 0.006). Consequently, higher AJCC stages were observed in RR-PTC patients (*P* = 0.001). Compared with patients in the control group, patients in the RR-PTC group received larger doses of I-131 after the initial resection of primary tumors. Notably, all six patients who died were in the refractory group.

#### 2.2 Molecular profile of PTCs correlated with I-131 resistance

A total of 23 genes were sequenced (Sup table S1). Four genes were found to harbor driver mutations (Table 2). The *BRAF*^V600E^ mutation was identified in 25 samples (56%), with no significant difference between the RR-PTC and the control groups (54.3% *vs.* 60.0%, *P* = 1.000). All six samples with *TERTp* mutations (17.1% *vs.* 0%, *P* = 0.312) were in the RR-PTC group. In addition, *RET* and *TP53* mutations were found in one and two patients, respectively. *RAS* mutations, including *HRAS*, *KRAS* and *NRAS*, were not detected in any samples. In addition, four kinds of gene fusions were found, namely, eight samples with *NCOA4*-*RET*, two with *CCDC6*-*RET*, three with *ETV6*-*NTRK3* and one with *STRN*-*ALK*. Fusions were more frequent in the control group (28.6% *vs.* 40.0%, *P* = 0.700), whereas *RET*-related fusions were almost equally distributed (22.9% *vs.* 20.0%, *P* = 1.000). However, no significant differences in the expression of these genes were found between the two groups.

**Table 2.** The comparison of gene mutations and fusions in RAI-refractory and RAI-sensitive PTC

| Comparison | BRAF | RET | TERTp | TP53 | Fusion <sup>a</sup> | RET fusion | NCOA4-RET |
| --- | --- | --- | --- | --- | --- | --- | --- |
| RR-PTC vs. Control |  |  |  |  |  |  |  |
| RR-PTC (35) | 19 (54.3) | 0 (0) | 6 (17.1) | 1 (2.9) | 10 (28.6) | 8 (22.9) | 7 (20.0) |
| Control (10) | 6 (60.0) | 1 (10.0) | 0 (0) | 1 (10.0) | 4 (40.0) | 2 (20.0) | 1 (10.0) |
| *P* value | 1.000 | 0.222 | 0.312 | 0.399 | 0.700 | 1.000 | 0.661 |
| Subgroups |  |  |  |  |  |  |  |
| ID (9) | **1 (11.1) \*** | 0 (0) | 1 (11.1) | 1 (11.1) | 4 (44.4) | 4 (44.4) | **4 (44.4) \*** |
| iDF (9) | 6 (66.7) | 0 (0) | **3 (33.3) \*** | 0 (0) | 1 (11.1) | 1 (11.1) | 0 (0) |
| iDG (17) | 12 (70.6) | 0 (0) | 2 (11.8) | 0 (0) | 5 (29.4) | 3 (17.6) | 3 (17.6) |
| Id (10) | 6 (60.0) | 1 (10.0) | 0 (0) | 1 (10.0) | 4 (40.0) | 2 (20.0) | 1 (10.0) |
| *P* value | 0.026 * | 0.311 | 0.193 | 0.411 | 0.421 | 0.327 | 0.081 |
| I-131 uptake |  |  |  |  |  |  |  |
| Negative (26) | 18 (69.2) | 0 (0) | 5 (19.2) | 0 (0) | 6 (23.1) | 4 (15.4) | 3 (11.5) |
| Positive (19) | 7 (36.8) | 1 (5.3) | 1 (5.3) | 2 (10.5) | 8 (42.1) | 6 (31.6) | 5 (26.3) |
| *P* value | 0.039 * | 0.422 | 0.222 | 0.173 | 0.206 | 0.281 | 0.253 |

\* $P < 0.05$ . Bold font indicated statistical significance revealed by $z$ test.
<sup>a</sup> Detected gene fusions included *ETV6-NTRK3* in three samples, one *STRN-ALK*, two *CCDC6-RET* and eight *NCOA4-RET*. The latter two fusions were collectively called *RET* fusion.

The forty-five samples were then divided into four subgroups. The *BRAF*^V600E^ mutation was less common in the continuous RAI uptake (ID) group (11.1%, *P* < 0.05), whereas the *TERTp* mutation was more frequent in the loss of uptake at the first I-131 treatment (iDF) group (33.3%, *P* < 0.05). Notably, *NCOA4-RET* fusion was more common in the ID group (44.4%, *P* < 0.05).

The present study further evaluated the associations between gene variants and I-131 uptake. The samples in the ID and RAI-sensitive PTC (Id) groups were categorized as positive for I-131 uptake. *BRAF*^V600E^ mutation was more frequent in the negative I-131 uptake group with statistical significance (69.2% *vs.* 37.8%, *P* = 0.039). The incidence of *TERTp* mutations was also greater in this group (19.2% *vs.* 5.3%, *P* = 0.222). In contrast, fusions were more common in the positive uptake group (23.1% *vs.* 42.1%, *P* = 0.206).

The distribution of gene variants with unknown clinical significance was also assessed (Sup table S2). The four mutations were located on *ATM* (c.1236-2A>T), *PTEN* (c.402G>A) and *RET* (c.2071G>A and c.2671T>G). Three variants were more common in the RR-PTC group, except for *PTEN* c.402G>A, although the difference was not statistically significant. In subgroup analysis, the incidence of *PTEN* c.402G>A was greater in the ID group (22.2%, *P* < 0.05). In addition, *PTEN* c.402G>A was associated with positive I-131 uptake with marginal significance (0% *vs.* 15.8%, *P* = 0.068).

#### 2.3 Overview of the proteomic profiling

A total of 9769 proteins were identified in 168 tissue slides from 73 patients. (Sup figure 1A). The principal component analysis (PCA) plot indicates no observable batch effect during data acquisition (Sup figure 1B). Tumor tissues mixed with little difference but negative lymph nodes were clustered (Sup figure 1C). The *R* values of correlation analyses exceeded 0.97 in both intra-batch and inter-batch tests, indicating high technical stability across the data acquisition (Sup figure 1D&E). Overall, no observable batch effect was detected (Sup figure 1F). Next, a filtering criterion was applied to select proteins with missing values less than 50%. These 7406 filtered proteins were then subjected to data normalization and outlier substitution (Sup figure 2A-C). After ID conversion, an expression profile encompassing 7394 proteins was compiled (Sup figure 2D). The median tumor purity was 76.2% (Sup figure 2E), indicating the reliability of our dataset for further analysis.

Subsequently, the original PTC lesions were clustered using dimensionality reduction based on the global proteomic profiles. The RAI-sensitive samples were more tightly clustered than the RR-PTC samples (Figure 1A). The overlap of the two groups might be explained by the similarity between tumor tissues and the intrinsic heterogeneity of RR-PTC samples. To minimize potential heterogeneity, RR-PTCs were furtherly divided into three subgroups (iDF, iDG and ID). The diversity of the ID group was the lowest among the three subgroups, while the diversity of the RAI uptake lost gradually (iDG) group was the highest (Figure 1B). Interestingly, when labeled with the *BRAF*^V600E^-mutation status, samples in the two groups were separately clustered (Figure 1C).

**Figure 1.**
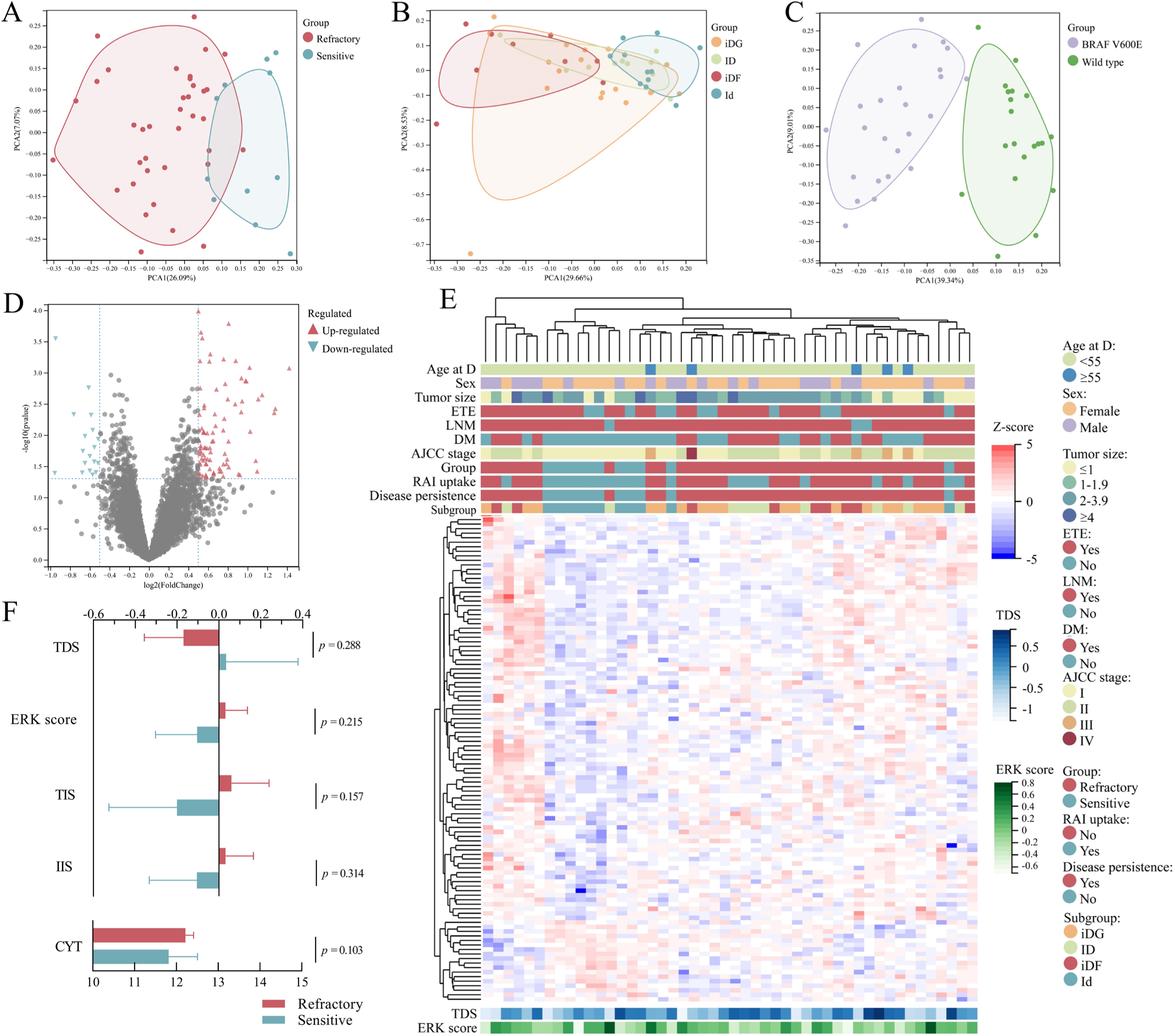
Proteomic overview of RAI-refractory and RAI-sensitive PTC samples. (A-C) Dimension reduction visualization of the analyzed PTC samples. The area covered by the samples represented similarity within a group. The samples were labeled according to their response to I-131 treatment (refractory or sensitive), subgroup (iDF, iDG, ID or Id) and *BRAF* mutation status. Three samples whose quality control for sequencing failed were not included in Figure C. (D) Differential expression analysis of samples in the RR-PTC and control groups. Unadjusted *P* values < 0.05 and absolute FC > 1.414 were defined as thresholds for DEPs. (E) Overview of the proteomic profiles. *Z* scores of the DEPs were calculated. (F) Correlation of scores and groups. Error bar denoted 95% CI. The TDS was lower in the RR-PTC group (−0.17 ± 0.562 *vs.* 0.03 ± 0.512, *P* = 0.288). Conversely, the ERK score was greater in the RR-PTC group (0.03 ± 0.317 *vs.* -0.10 ± 0.297, *P* = 0.215). The means and SDs are presented with boxes and error bars. Abbreviations: age at D, age at diagnosis; DM, distant metastasis; ETE, extrathyroidal extension; LNM, lymph node metastasis.

A total of 107 differentially expressed proteins (DEPs) with *P* values < 0.05 and absolute fold change (FC) > 1.414 were identified (Figure 1D). Eighty-nine proteins were upregulated in the RR-PTCs, whereas the remaining eighteen proteins were downregulated. The top upregulated protein in the RR-PTCs was KRT71 (log_2_FC = 1.422). Interestingly, the top downregulated protein also belonged to the keratin family (KRT16, log_2_FC = -0.955). Samples were clustered unsupervisely based on the 107 DEPs and then labeled with clinicopathological features (Figure 1E). The borders of the clinicopathological labels were generally indistinct. However, the thyroid differentiation score (TDS) and extracellular signal-regulated kinase (ERK) score were correlated with the groups to some extent (Figure 1F). The TDS was lower in the RR-PTC group, which could indicate poorer differentiation (*P* = 0.288). Conversely, the ERK score was greater in the RR-PTC group (*P* = 0.215). Unfortunately, none of these scores reached statistical significance.

Gene set enrichment analysis (GSEA) revealed 64 pathways with statistical significance (Sup figure 3A). Twenty-nine pathways with positive normalized enrichment scores (ESs) were upregulated in RR-PTC samples. The pathway with the greatest increase was gamma carboxylation hypusine formation and arylsulfatase activation (nES = 1.687). In addition, Akt phosphorylates targets in the cytosol was also upregulated (nES = 1.552). Notably, *RAS* processing was downregulated (nES = -1.728). The Kyoto Encyclopedia of Genes and Genomes (KEGG) database was also used for the annotation of pathways closely related to oncogenesis and dedifferentiation (Sup figure 3B). P53 signaling was significantly enriched (nES = 1.5440, *P* = 0.0136). Interestingly, the MAPK signaling pathway showed a slight positive correlation with RR-PTCs (nES = 1.2623, *P* = 0.1642). The gene set variation analysis (GSVA) results supported the above results (Sup figure 3C).

A previously published study assessed the transcript profiles of RR-PTC via a gene microarray (21). The DEPs identified in the present study were assessed via this profile. Only three DEPs were dysregulated (Sup figure 3D). BET1 and C11orf96 were downregulated in the RR-PTC group, whereas SLC4A9 was greater. Notably, the regulation direction of BET1 and SLC4A9 was consistent with the results of the present study, but C11orf96 was oppositely expressed in the two analyses. GSVA revealed two differentially downregulated pathways, none of which were enriched in our study (Sup figure 3E&F).

To assess the spectral change in RR-PTC, the Id, ID and iDF subgroups were defined as having low, intermediate and high severity, respectively. A total of 403 proteins with significance difference were divided into six clusters (Sup figure 4). In two clusters, 160 proteins were monotonically regulated (Sup figure 5A). Collagen biosynthesis and modifying enzymes were significantly enriched (Sup figure 5B&C). Regulation of protein autophosphorylation and regulation of extrinsic apoptotic signaling pathway were also associated with the severity of RR-PTC.

#### 2.4 Proteomic biomarkers and pathways associated with I-131 uptake

For analysis of proteomic biomarkers and the underlying mechanisms for the loss of I-131 uptake in PTC, two subgroups, ID and iDF, were included in the subsequent analysis. The ability of these two groups to take up I-131 was completely different at the first radiotherapy. The demographic and clinicopathologic characteristics of the two groups were roughly comparable (Sup table S3). Since the diagnosis of RR-PTC in the ID group required multiple rounds of I-131 treatment, the cumulative dose of I-131 was inevitably greater than that in the iDF group (*P* = 0.001).

Although the ID and iDF groups overlapped with each other to some extent, a total of 145 DEPs were identified (Figure 2A&B). Ninety-five proteins were upregulated in the ID group, whereas the remaining 50 proteins were highly expressed in the iDF group. The 145 DEPs could barely separate the two groups. The TDS was greater in the ID group, whereas the ERK score was lower. However, none of these scores reached statistical significance (Figure 2C&D).

**Figure 2.**
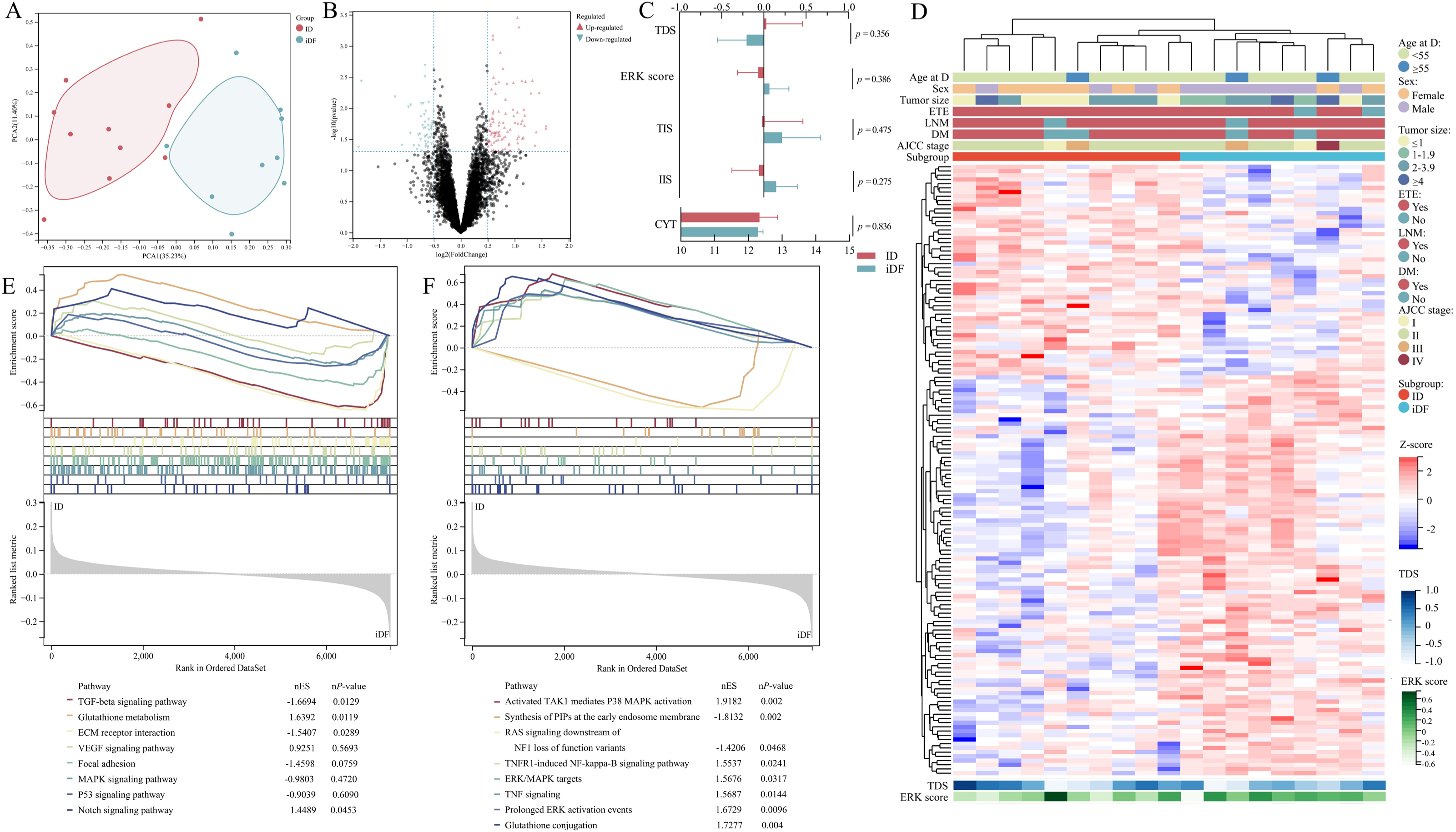
Differential expression analysis of the ID and iDF groups. (A) PCA of the two groups. (B) Differential expression analysis of samples in the two groups. Unadjusted *P* values < 0.05 and absolute FC > 1.414 were defined as thresholds for DEPs. (C) Correlations of scores and groups. Error bar denoted 95% CI. The TDS was greater in the ID group (0.03 ± 0.599 *vs.* -0.20 ± 0.454, *P* = 0.356), while the ERK score was greater in the iDF group (−0.07 ± 0.345 *vs.* 0.07 ± 0.300, *P* = 0.386). (D) Expression profiles of 145 DEPs. (E&F) GSEA based on the KEGG and Reactome databases. Sixteen pathways are presented in the plot.

Pathway enrichment analysis was then conducted. Only seven pathways in the KEGG database were significantly enriched. Among the seven pathways, glutathione metabolism and Notch signaling were enriched in the ID group whereas TGF-beta signaling and ECM receptor interaction were enriched in the iDF group (Figure 2E). Fifty-one pathways in the Reactome database were significantly enriched. The top enriched pathway was “activated TAK1 mediates P38 MAPK activation” (Figure 2F). In addition, TNF signaling, glutathione conjugation and several MAPK-related pathways, including ERK/MAPK targets and prolonged ERK activation events, were correlated with positive I-131 uptake.

The results of pathway enrichment were then assessed via GSVA. Sixty-three pathways were significantly enriched and 18 of these pathways overlapped with the GSEA results (Figure 3A). After the associations between pathways and tumor progression were evaluated, sixteen pathways were found to play roles in the loss of I-131 uptake in PTC, eleven of which overlapped with the GSEA results (Figure 3B). Six pathways were closely related to MAPK signaling. Three pathways are associated with transforming growth factor-β activated kinase 1 (TAK1) signaling. In addition, one pathway was correlated with both MAPK and TAK1 signaling. The other six pathways involved glutathione metabolism, TNF signaling and MET proto-oncogene (MET) signaling. Most proteins in these pathways were not dysregulated (Figure 3C).

**Figure 3.**
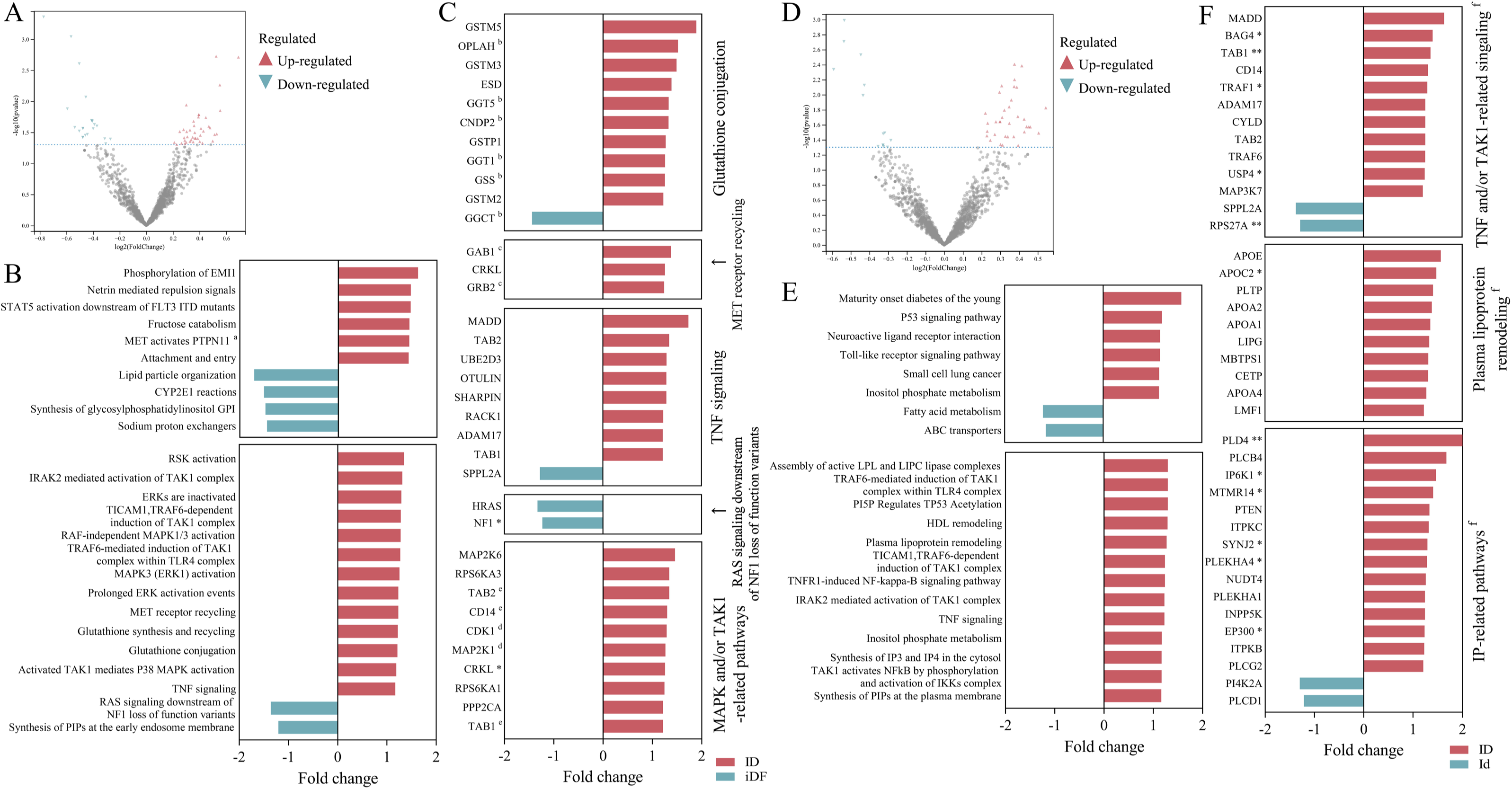
Expression of selected genes according to the enriched pathways identified in the ID *vs.* iDF and ID *vs.* Id comparisons. (A&D) Dysregulated pathways identified in the ID *vs.* iDF and ID *vs.* Id comparisons. The *P* value threshold was 0.05. No threshold was set for fold change. GSVA was conducted based on the Reactome annotation file. (B) The ten most significantly enriched pathways and the thirteen selected pathways. (C) The selected proteins from the thirteen selected pathways. The expression of proteins involved in these thirteen pathways was compared between the RAI-positive and RAI-negative groups. Proteins with an absolute FC > 1.2 were displayed. (E) The eight significantly enriched pathways and thirteen selected pathways. (F) The selected proteins from the thirteen selected pathways. The TNF signaling pathway and the TAK1-related pathways were combined in a single plot since the proteins strongly overlapped. * *P* < 0.05, ** *P* < 0.01. ^a^ The pathway “MET activates PTPN1” was also selected for subsequent identification of proteins. ^b, c^ Involved in the selected pathways “glutathione synthesis and recycling” and “MET activates PTPN11”, respectively. ^d^ Both CDK1 and MAP2K1 participate in the pathways “MAPK3 (ERK1) activation” and “RAF-independent MAPK1/3 activation”, and MAP2K1 is also a participant in “prolonged ERK activation events”. ^e^ TAB2 and TAB1 are involved in both MAPK and TAK1-associated pathways, while CD14 participates in TAK1-associated pathways. ^f^ Molecules in these figures might participate in several pathways. The associations of molecules and pathways are provided in Sup table S5.

The results were validated with a transcriptomic profile of RR-PTCs (GSE151179). The samples in GSE151179 were categorized as ID or iD based on their I-131 uptake. The iD group was not subdivided into iDF or iDG since the information was not provided. Three common differentially expressed genes (DEGs) were identified (Sup figure 6A). However, the regulatory direction was completely different. GSVA revealed five common pathways (Sup figure 6B), and the direction of the change was consistent between the two profiles. Notably, three TAK1-associated pathways were significantly enriched. All genes involved in the 16 selected pathways and these five common pathways were tested. Only three genes were differentially expressed between the two groups (Sup figure 6C).

Immune analysis revealed an abundancy of memory B cell and a deficiency of macrophage and immature T cell in the ID samples (Sup figure 7A). Notably, eosinophils were more abundant in RAI-sensitive samples compared to RR-PTCs. However, immunohistochemistry results did not show any statistically significant differences (Sup figure 7B).

#### 2.5 Proteomic biomarkers and pathways for different responses to positive I-131 uptake

The data of twenty-one patients in the ID group and the Id group (or control group) were compared to determine the potential mechanisms underlying the different responses to positive I-131 uptake in PTC patients. No significant differences in demographic or tumor features were found, except for a greater incidence of ETE in the ID group (*P* = 0.035) (Sup table S4). More distant metastases were found in the ID group (80% *vs.* 9.1%, *P* = 0.002), and consequently, the disease stages were greater (*P* < 0.001). According to the criteria of the ID group, the cumulative I-131 dose was inevitably greater (*P* < 0.001).

The two groups could be distinctly separated from each other (Figure 4A). Twenty-one samples were roughly divided into two groups with 125 DEPs identified (Figure 4B&D). No significant differences were found in the TDS, ERK score, T cell infiltration score (TIS), immune infiltration score (IIS) or immune cytolytic activity score (CYT) (Figure 4C).

**Figure 4.**
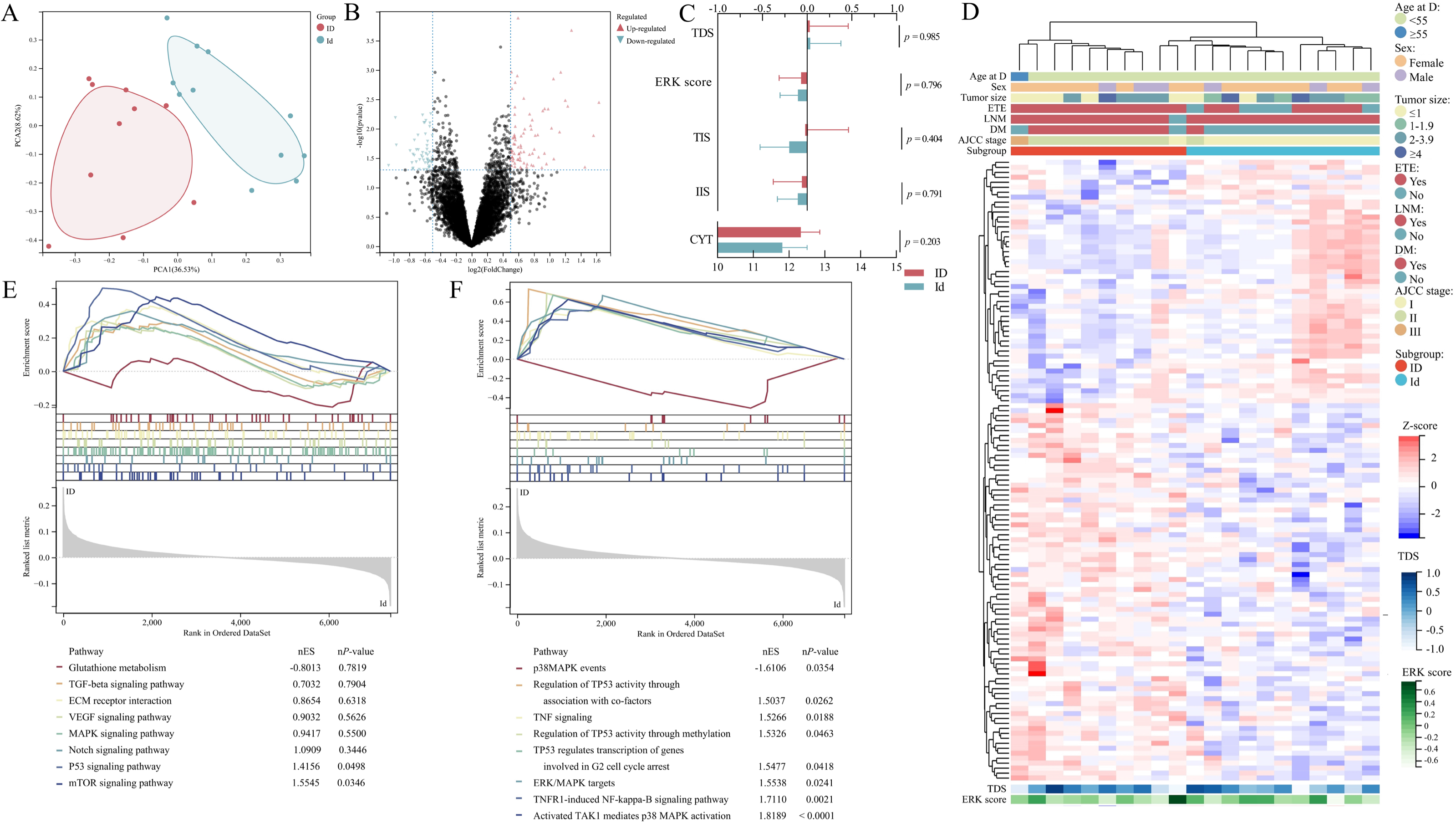
Differential expression analysis of the ID and Id groups. (A) PCA of the two groups. (B) Differential expression analysis of samples in the two groups. Unadjusted *P* values < 0.05 and absolute FC > 1.414 were defined as thresholds for DEPs. (C) Correlations of scores and groups. Error bar denoted 95% CI. No significant difference was found in the TDS (0.03 ± 0.599 *vs.* 0.03 ± 0.512, *P* = 0.985) or ERK score (−0.07 ± 0.345 *vs.* -0.10 ± 0.297, *P* = 0.796). (D) Expression profiles of 125 DEPs. (E&F) GSEA based on the KEGG and Reactome databases. Sixteen pathways are presented in the plot.

GSEA revealed twelve enriched pathways based on KEGG annotations. The P53 signaling pathway and mTOR signaling pathway were both significantly upregulated in the RAI-resistant samples (Figure 4E). Interestingly, inositol phosphate (IP) metabolism was also activated in the ID group (*P* = 0.0190). No significant difference was revealed in the MAPK signaling pathway. Forty-five pathways were significantly enriched according to the Reactome annotation file. TP53-related pathways, including regulation of TP53 through association with co-factors (*P* = 0.0260) and methylation (*P* = 0.0463), were increased in ID samples (Figure 4F). TNF signaling (*P* = 0.0188) and its participant TNFR1-induced NF-kappa-B signaling pathway (*P* = 0.0021) were both associated with RAI uptake and resistance. In addition, the pathway “activated TAK1 mediates p38 MAPK activation” was closely related to RAI-resistance (*P* < 0.0001). An increase in IP metabolism was also detected (*P* = 0.0081).

GSVA was conducted to verify the previous results. A total of 49 pathways were enriched with significant differences, eleven of which overlapped with previous results (Figure 3D). After the search and selection, four clusters comprising thirteen pathways were pooled for subsequent analysis (Figure 3E). Eight of the thirteen pathways were also dysregulated according to GSEA. Thirty-nine proteins involved in TNF signaling, TAKI-related pathways, plasma lipoprotein remodeling and IP-related pathways were identified with an absolute FC > 1.2 (Figure 3F).

The expression levels in the transcriptomic profiles of the ID and Id groups were then compared for validation. Only PGS1 was downregulated in the ID group (Sup figure 6D), which was consistent with the proteomic data. According to the pathway analysis, only GAB1 signalosome was significantly decreased in the ID group (Sup figure 6E). However, none of the genes in the GAB1 signalosome were differentially expressed between the two groups. Three genes in the above thirteen pathways were downregulated in the ID group, in contrast to the expression pattern in the proteomic profile (Sup figure 6F).

#### 2.6 Association of gene variance and protein expression

The 7394 proteins were divided into 21 modules (Figure 5A&B). *BRAF* mutation was significantly correlated with seven gene modules, among which the blue module had the highest absolute correlation coefficient (CC) (*P* = 8.4e-6). The most prominent module associated with *TERTp* mutations was pink (*P* = 1.9e-3). The green-yellow module was closely related to the gene fusions (*P* = 3.7e-3) and the *NCOA4-RET* fusion (*P* = 0.03). For gene variants of unknown significance, *ATM* c.1236-2A>T and *RET* c.2671T>G was related to the green and yellow module, respectively (*P* < 0.01).

**Figure 5.**
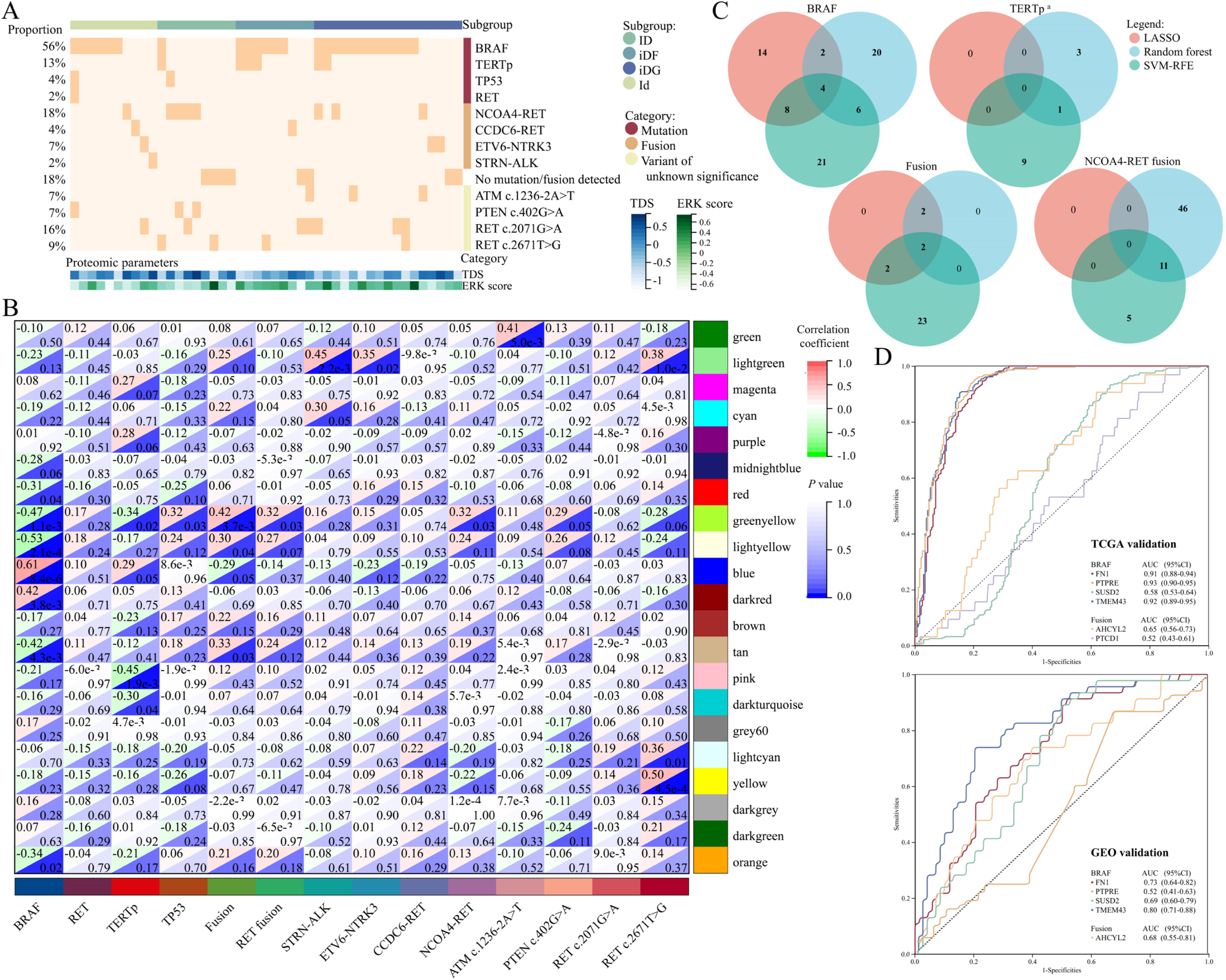
Gene variants and WGCNA. (A) Gene variance landscape. (B) Correlations of gene modules and gene phenotypes. The cells were divided into upper left (correlation coefficient) and lower right (*P* value) parts. (C) The intersection of LASSO-, RF- and SVM-RFE-identified biomarkers for *BRAF*, and *TERTp* mutations, gene fusions and *NCOA4-RET* fusion. (D) Validation of six genes in the TCGA-THCA cohort and merged GEO cohort. PTCD1 was not detected in the merged GEO dataset. ^a^ The low number of *TERTp*-positive samples (n = 6) prevented model construction and error analysis for *TERTp* mutations via the SVM algorithm. The first ten proteins with the top average rank were processed for biomarker identification.

Biomarker proteins were then selected from the most correlated modules of each gene mutation and fusion. Since other gene mutations were generally rare (< 10%) in the present study, samples with *BRAF* mutation, *TERTp* mutation, gene fusion and *NCOA4-RET* fusion were retained for subsequent analysis. A total of 28, 32 and 39 proteins were used for model construction of *BRAF* mutations via three machine learning algorithms (LASSO, RF and SVM-RFE), and four of these proteins were common biomarkers (Figure 5C). In addition, only PTCD1 and AHCYL2 were revealed to be common biomarkers for gene fusions. According to the receiver operating characteristic (ROC) curves of The Cancer Genome Atlas Thyroid Cancer (TCGA-THCA) dataset, the areas under the curve (AUCs) of three genes exceeded 0.9 (Figure 5D). PTPRE had the highest accuracy (AUC = 0.93), followed by TMEM43 (AUC = 0.92) and FN1 (AUC = 0.91). According to the merged Gene Expression Omnibus (GEO) profile, the most accurate gene was TMEM43 (AUC = 0.80), followed by FN1 (AUC = 0.73). Interestingly, PTPRE did not significantly differ. The AUCs of AHCYL2 for gene fusions in TCGA-THCA and merged GEO cohorts were 0.65 and 0.68, respectively. Disease free survival analysis revealed that dysregulation of FN1 (*P* = 0.026) and AHCYL2 (*P* = 0.038) was associated with poor prognosis in patients with thyroid cancer (Sup figure 8).

To explore the potential pathways in case of *BRAF* mutation and wild-type *TERTp*, nineteen *BRAF*-mutated and *TERTp*-unmutated samples were analyzed. A total of 184 proteins were significantly different between the RAIR and control groups (Sup figure 9A&B). Several pathways, which were related to the immune response, including T cell activation, regulation of lymphocyte activation and phagocytosis, were enriched with statistical significance (Sup figure 9C). Granzyme A signaling was downregulated in *BRAF*-wild-type samples, whereas neutrophil degranulation, B cell development and phagosome formation were upregulated in *BRAF*-mutated *TERTp*-unmutated samples (Sup figure 9D).

### 3 Discussion

To our knowledge, the present study is the first to construct a proteomic profile of RR-PTC on primary lesions. We evaluated the differences in the expression of genes in the RAI-refractory and RAI-sensitive samples. Three genes were potential biomarkers for RR-PTC. To minimize the intrinsic heterogeneity due to different criteria, we subdivided RR-PTC primary lesions based on the response to radiotherapy and the presence of I-131 uptake. MAPK signaling was the most commonly involved pathway. TGF-beta and TAK1 signaling was revealed to be associated with the loss of I-131 uptake. In contrast, tumor necrosis factor (TNF) signaling might affect cancer cell death in positive foci. Targeted deep sequencing revealed that wild-type *BRAF* and *TERTp* mutation were associated with the ID and iDF phenotypes, respectively. *NCOA4-RET* fusion was more frequent in the ID group. With respect to protein expression and gene variance, four proteins were associated with *BRAF*^V600E^ mutation.

The associations between thyroid cancer and gene variants have been widely discussed. The pathway most likely related to dedifferentiation and RAI refractoriness was MAPK signaling (22). MAPK signaling regulates cell proliferation, dedifferentiation and death. *BRAF* is an essential kinase in the cascade. The *BRAF*^V600E^ mutation has been reported to be associated with aggressive tumor behavior and poor prognosis (23, 24). Recent studies have revealed a strong correlation between *BRAF*^V600E^ mutation and negative iodine uptake (12, 21), which was confirmed by a meta-analysis (25). The overall prevalence of *BRAF* mutation was ∼55% in our study, but ID samples were less likely to harbor *BRAF* mutation (∼10%) than other subgroups. Given that *BRAF* mutation can impair the expression of the NIS protein (26), recurrent or metastatic lesions without *BRAF* mutation are prone to be positive in I-131 WBSs. Interestingly, a more recent study by Mu *et al.* revealed a lower rate of *BRAF* mutation in the “continuously RAI-avid but RAI-refractory” group (∼14%) than in the partial and gradual RAI-refractory groups (27). Although few studies have subdivided RR-PTC based on different criteria, we could hypothesize that ID tumors have distinct molecular features and tumorigenic mechanisms.

However, it is difficult to explain the high prevalence of *BRAF* mutation in the Id group in our study, which was not consistent with many previously published articles (27, 28). Some studies have reported no significant correlation between *BRAF* status and RAI uptake (29, 30). It was assumed that clinical pathways in different countries/regions and patient selection protocols contributed to the diverse observations to some extent (24).

In addition, *TERTp* mutation is a promising predictive factor for the RAIR. The prevalence of this mutation was lower than that of the *BRAF* mutation, but patients harboring *TERTp* mutations had poorer prognoses and greater mortality (31). Liu *et al.* reported that the genetic duet of *BRAF* and *TERTp* mutations robustly predicted the loss of RAI avidity in PTCs with a high positive predictive value (97.4%) (12). However, the negative predictive value was lower than 50%, which limits its utility in clinical practice. Subsequent studies have shown similar but somewhat different results (13). The genetic duet could predict RAIR with a positive predictive value barely higher than 50%. A recent study from Shanghai revealed that *TERT* accelerated *BRAF*-mutated thyroid cancer dedifferentiation and progression by regulating ribosome biogenesis (32). Our study also revealed a greater rate of *TERTp* mutation in the RR-PTC group. Interestingly, *TERTp* mutation was significantly more frequent in iDF samples, indicating its strong association with the loss of RAI uptake. This observation was in accordance with a previously published article (27). Since all six *TERTp*-mutated samples harbored the *BRAF*^V600E^ mutation, we did not test the combined efficacy of the two mutations.

Of interest, our study revealed a significantly greater incidence of *NCOA4-RET* (*PTC3-RET*) fusion in the ID group. This phenomenon was also observed in two studies. An Italian study revealed that fusion genes (especially *RET-PTC*) were more common in ID (39%) than in iD (16%) and Id (13%) patients (*P* = 0.075) (21). Mu’s study revealed that the incidence of *RET*-fusions in the ID subgroup (∼40%) was greater than that in other RR-DTC subgroups and the Id group (27). However, statistical significance was not reached in the above two studies. A subsequent study from Mu’s institution revealed that fusion oncogenes in pediatric DTCs were associated with RAI-refractoriness (*P* = 0.017) (33). But *RET*-fusions were not analyzed separately. The radioactive isotope is concentrated in thyroid epithelium-derived tumor cells and exerts tumor-killing effect by emitting β-rays. Considering that *RET* rearrangements can activate its kinase and inhibit apoptosis via the MAPK and PI3K signaling pathways (34), the apoptosis triggered by radioisotope-induced DNA damage could also be impaired by *RET* alterations, especially the *NCOA4-RET* fusion. The potential causal relationship between *RET* fusion and the ID phenotype has not been verified in cell or animal experiments. Recent clinical studies have shown that the *RET* fusion-directed therapy can restore RAI avidity in patients with RR-PTC (35, 36).

The accuracy of gene mutations and fusions are helpful but not satisfactory. Proteomic analysis was conducted to identify promising biomarkers related to the RAIR. Although the expression differences between phenotypes were generally not obvious, some signaling pathways were significantly enriched. In the comparison of the RAI-sensitive and RAI-refractory groups, P53 signaling was the pathway most likely associated with the loss of iodine uptake. Although some scholars have suggested that *TP53*-mutated follicular adenomas are precursors for the dedifferentiation of anaplastic thyroid cancer (37), few studies have investigated the correlation between the RAIR and mutations in P53 signaling. At that time, we assumed that the heterogeneity in RR-PTC reduced the power of the test. In the subsequent comparisons of subgroups (ID *vs.* iDF and ID *vs.* Id), enriched pathways clustered into several major pathways, including the MAPK signaling and TNF signaling pathways. The MAPK pathway is strongly related to thyroid cancer behaviors, and *BRAF* mutation impairs the expression of the NIS protein by deacetylating its gene promoter histones (26). MAPK inhibitors and histone deacetylase inhibitors can restore iodine uptake and re-differentiate PTC cells (38, 39). Interestingly, our study revealed that TAK1-associated pathways were also correlated with that RAIR. A recent study suggested that silencing TAK1 inhibited the proliferation and migration of thyroid cancer cells via that suppression of p38 MAPK signaling (40). We hypothesize that TAK1 is a novel potential molecular target of RAIR. The TNF signaling is another pathway of interest. TNF is an important cytokine that triggers inflammation. A retrospective study by Gheorghe *et al.* suggested that TNF-α might exert different antitumor effects in response to RAI therapy depending on the patient’s immune profile (41). The activation of TNF signaling might restore I-131 uptake and promote redifferentiation in thyroid cancer cells. Unfortunately, most proteins involved in these pathways did not significantly differ.

Several previous studies have plotted the molecular and omic atlas of RR-DTC (Sup table S6). Sabra and Shobab independently constructed the genomic landscape for RR-DTC (42, 43). Mutations in the MAPK signaling pathway accounted for ∼50% of oncogenic drivers, which was similar with our results. In 2020, Colombo *et al.* constructed molecular and gene/miRNA profiles for RR-PTC (21). *BRAF* mutation was more frequent in the RAI-negative RR-PTC group. Although PTCs were clearly distinguished from normal thyroid tissues, no distinct expression patterns were found between RAI-refractory and RAI-sensitive PTCs. In general, our results were consistent with those of Colombo’s study. A distinguishable feature for RAIR was not found based on either the mRNA/miRNA or protein profile, regardless of subclassification. The insufficient sample size may partly explain this phenomenon. One hypothesis is that the RAIR occurs during recurrence and metastasis. The hypothesis, however, is not convincing in the presence of some RAI negative lesions at the first I-131 treatment. Another competing hypothesis is that the underlying mechanisms of the RAIR are trivial and are covered by the heterogeneity of other tumor behaviors. More homogenous samples and proper subclassification are needed to eliminate confounding effects and uncover the underlying mechanisms involved. Two precursor studies provided novel insights into the proteomic expression of RR-PTC but were limited by small sample sizes and the lack of primary tumors (44, 45).

The current study has several shortcomings. First, although the medical records of more than ten thousand patients were reviewed thoroughly by independent researchers, the intrinsic nature of this single-center, retrospective study and the relatively small sample size limit the generalizability of our conclusions. In addition, the lack of Benjamini-Hochberg correction for differential expression analysis undermined the reliability of the results. In addition, due to the lack of previous proteomic profiles, potential protein biomarkers were tested with several external transcriptomic profiles. The consistency of expression across mRNAs and corresponding proteins is debatable (20). Moreover, FTC samples and metastatic lesions were excluded from the present study because of the small sample size and potential histopathological heterogeneity. The potential biomarkers and mechanisms were not verified with *in vivo* experiments. The above observations of our study could be confirmed with multicenter large-sample proteomic profiling. Cell and animal experiments could help verify our results.

In conclusion, a proteomic profile based on RR-PTC primary lesions was constructed for the first time in the present study. Our current work improves the molecular and biological understanding of RR-PTC, which could enlighten future preclinical and clinical studies toward molecule-guided treatment. The dataset created in this study could serve as an important resource for further investigations of RR-PTC biology and therapeutic targets.

### 4 Patients and Methods

#### 4.1 Study design and ethics approval

This retrospective study was conducted at Renmin Hospital of Wuhan University. The Institutional Ethical Committee of the hospital reviewed and approved the study design (No. WDRY2021-K032). The requirement for obtaining informed consent from the involved patients was waived due to the retrospective nature of the study. The study was conducted in accordance with the Declaration of Helsinki (46). The study was performed in accordance with the STROBE checklist for case-control study (version 4).

#### 4.2 Patient and tissue selection

The medical records of patients who were diagnosed with thyroid cancer at our tertiary center from Jan. 1^st^, 2016 to Dec. 30^th^, 2022 were reviewed (Figure 6A). Patient demographic and clinicopathological characteristics, laboratory test results and radiological results were collected by two researchers and independently evaluated. During the initial screening, patients who a) had incomplete medical records and b) did not undergo I-131 WBS were excluded. In the second round of assessment, histopathological data and radiological images, including chest computed tomography (CT), cervical magnetic resonance imaging (MRI), whole-body bone scanning and positron emission tomography (PET), were used to evaluate local and distant metastasis at the first admission for surgery. Patients without cervical or distant metastasis at diagnosis were excluded from the subsequent evaluation. Patients with RAI uptake limited to thyroid bed and disease remission were also excluded. In the final assessment, RAI uptake was determined by I-131 WBSs or I-131 single photon emission computerized tomography (SPECT). A total of 73 patients were identified to have locoregional or distant metastasis, which was confirmed by radiological, cytopathological and/or histopathological solid evidence. Patients with primary lesions not resected in our center or other histopathological types other than PTC were also excluded from further statistical analysis. Finally, forty-eight PTC patients with locoregional and/or distant metastasis were included in this study and their primary tumor samples (one sample from each patient) were used for subsequent analysis.

**Figure 6.**
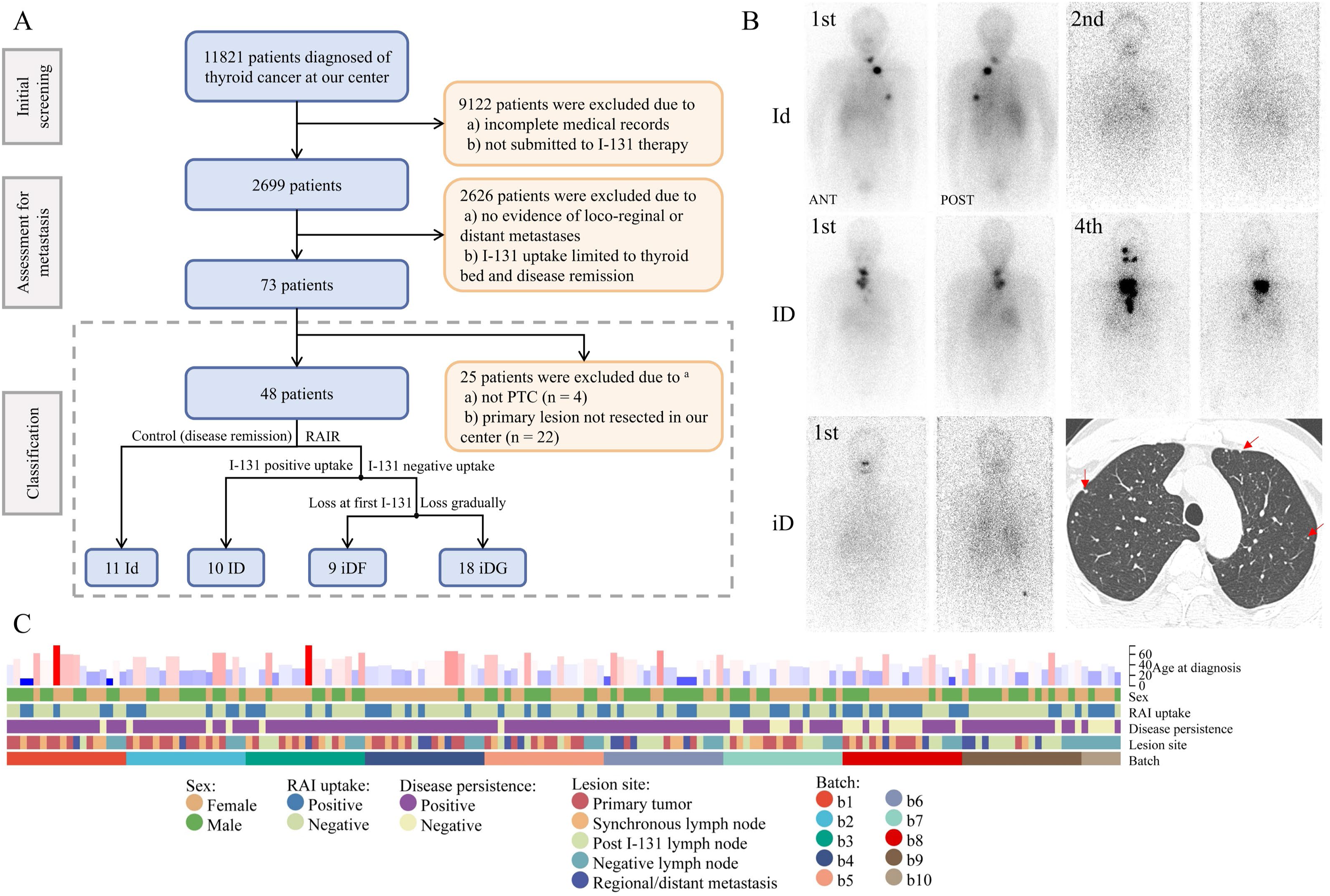
(A) Flow chart of patient and sample selection. Strict criteria were applied to identify eligible patients. Individuals with insufficient radiological evidence were excluded from the second round of selection. A total of 73 patients (in the dashed box) had regional or distant metastatic lesions with solid evidence. The FFPE tissues were collected for subsequent proteomic analysis and/or TNGS. Notably, 25 patients were excluded from this study due to a lack of primary lesions or not having PTC. (B) Grouping of included patients. Three samples are shown here for the Id, ID and iD groups. The first patient belonged to the Id group. The patient’s s lung metastases, which were confirmed by I-131 WBS and chest CT, were eliminated by two I-131 treatments. The second patient was included in the ID group. The patient received total thyroidectomy but the cervical regional metastatic lesion progressed during the four rounds of I-131 treatment. The first and second I-131 WBSs of the third patient revealed no positive focus. However, PET/CT during the same period revealed several suspicious pulmonary nodules (red arrow), which were confirmed to be metastases by subsequent surgery and histopathology. (C) Information on the collected FFPE samples. The protein profiles of 168 samples were evaluated in 10 batches. The sample types included primary lesions, synchronous and post I-131 cervical LNM, negative cervical lymph node and regional or distant metastatic lesions. Notably, the regional metastatic lesions were tumors near the primary site, such as the esophagus, trachea and cervical skin. LNM was not included in regional metastatic lesions. In addition, forty-eight primary tumors were examined via TNGS. ^a^ The primary lesion of one patient was neither resected in our center nor examined to be PTC.

The included patients were then divided into two groups based on their response to RAI treatment: RAI-sensitive (RAI uptake positive and disease remission, Id) and RAI-refractory (disease persistence, Figure 6B). RAI refractoriness was identified in accordance with the 2015 American Thyroid Association (ATA) guideline (7). Briefly, patients with metastatic lesions that a) did not ever concentrate RAI, b) lost the ability to concentrate RAI after previous evidence of RAI avidity, and c) concentrated in some lesions but not in others were considered to have negative RAI uptake. Individuals with d) metastatic disease despite a significant concentration of RAI were defined as having positive uptake. Owing to the potential heterogeneity of RR-PTC, patients in the RAI-refractory group were further categorized into three subgroups: a) negative RAI uptake at the first RAI treatment with disease persistence (iDF), b) RAI uptake lost gradually after previous RAI treatments with disease persistence (iDG), and c) positive RAI uptake but with disease persistence (ID). Tumor stage and the risk of recurrence were stratified using the American Joint Committee on Cancer (AJCC) staging system and the 2015 ATA guideline (7, 47).

Resected primary and metastatic lesions were preserved in formalin-fixed paraffin-embedded (FFPE) tissue blocks at room temperature. A total of 168 FFPE samples were collected from the above 73 patients (Figure 6C). The tissue types included primary tumors, synchronous cervical LNM, LNM after I-131 treatment, negative lymph nodes, and regional and distant metastatic lesions. Hematoxylin and eosin-stained slides from all samples were reviewed by two independent pathologists with expertise in thyroid pathology. The tissue blocks with the highest tumor purity in each patient were selected for subsequent analysis. The borderlines of the tumor and adjacent tissues were carefully marked by expert pathologists.

#### 4.3 Proteomic data acquisition and preprocessing

The FFPE samples were prepared for subsequent proteomic analysis as described previously (19, 48). The samples were allocated into ten batches. The tissues were subjected to a series of manipulations, including dewaxing, rehydration and lysis, for peptide extraction and digestion via pressure cycling technology (PCT). Peptides were then quantified via a liquid chromatography (LC) system coupled with a trapped ion mobility spectrometry mass spectrometer (MS). Data-independent acquisition (DIA) files were acquired and then analyzed against a thyroid tissue specific spectral library (19) using DIA-NN (v1.8.1) (49). Correlation analysis was conducted to assess the inter-batch and intra-batch stability of data acquisition. The profile was then processed for protein filtration, imputation and ID conversion. A detailed description of proteomic data acquisition, quality control and preprocessing can be found in the supplementary methods.

#### 4.4 Targeted next-generation sequencing (TNGS)

The primary lesions from 48 patients with PTC were subjected to molecular profiling via TNGS. The protocol was performed as described previously (50). Mutations and fusions were evaluated for 23 genes (Sup table S1). A detailed description of THGS can be found in the supplementary methods.

#### 4.5 Bioinformatic analysis

DEPs were identified using a multiple linear regression algorithm and analysis of variance (ANOVA). Differential expression analysis was conducted using the *limma* R package. Proteins with an absolute FC > 1.414 and a *P* value < 0.05 were defined as DEPs. The dimension of the profile was reduced and visualized using the PCA algorithm. ANOVA was used to identify proteins correlated with the severity of RR-PTC. Proteins with statistical significance were clustered using the *mfuzz* R package. Proteins in monotonically regulated clusters were regarded as DEPs.

Pathway enrichment analysis and gene ontology (GO) were conducted using four tools. GSEA was conducted using GSEA software to evaluate potential pathways and molecular mechanisms. For GSVA, ESs were calculated with the *GSEA* R package. Predefined gene sets were downloaded from the Reactome and KEGG databases. The top enriched GO processes were identified via the Metascape web-based platform. Another network tool, IPA, identifies most significantly relevant pathways with the overall activation or inhibition states based on DEPs.

Immune infiltration analysis was performed via the *CIBERSORTx*, *ESTIMATE* and *single sample GSEA* (*ssGSEA*) algorithms. Several scores, including the TDS, ERK score and CYT, were calculated based on the normalized protein profile. TIS and IIS were calculated using the Z score-standardized ssGSEA matrix. *BRAF*-*RAS* score (BRS) was not calculated since no *RAS*-mutated samples were identified.

Weighted correlation network analysis (WGCNA) was performed to cluster genes with high correlation and assess the correlation between protein modules and gene alterations. Proteins in the modules that were highly associated with gene mutations or fusions were selected. Least absolute shrinkage and selection operator (LASSO) regression, random forest (RF), and support vector machine-recursive feature elimination (SVM-RFE) were used to identify biomarker proteins based on selected proteins.

Several external datasets were used for the validation of our results (Sup Table S7). GSE151179 from the GEO database included 52 samples derived from radioiodine-refractory and radioiodine-avid PTC patients. This dataset was used to verify the results related to RAI refractoriness. The associations between gene alterations and protein expression were assessed with the THCA (thyroid cancer) program from TCGA and four additional datasets from the GEO database. The predictive performance of the genes in the external datasets was evaluated with ROC curves.

A detailed description of the bioinformatic analysis and references to methodological articles can be found in the supplementary methods.

#### 4.6 Statistical analysis

Quantitative variables were displayed as the means ± standard deviations or medians ± quartiles, whereas qualitative variables were presented as numbers and ratios. The significant differences in the quantitative variables were determined via two-tailed independent *t* test or Mann-Whitney *U* test, as appropriate. The *chi*-square test was used to evaluate the differences in the distributions of qualitative variables. When multiple groups were present, the *z* test with a Bonferroni correction was used to assess the intergroup difference in every group. Statistical analysis was conducted using SPSS software (IBM, US, v26).

## Disclosure

### Ethics approval statement

The Institutional Ethical Committee of the hospital reviewed and approved the study design (No. WDRY2021-K032). The requirement for obtaining informed consent from the involved patients was waived due to the retrospective nature of the study. The study was conducted in accordance with the Declaration of Helsinki.

## Supporting information

Supplementary methods, Supplementary figure 1-12, and References

## Data Availability

The proteomic data have been deposited in the iProX database with the project ID IPX0009103001. Calculation files and additional data are available in the Mendeley database (DOI: 10.17632/yfpfvktrxn). No custom code was used in the current study.

https://www.iprox.cn//page/SCV017.html?query=IPX0009103001

https://data.mendeley.com/datasets/yfpfvktrxn/3

## Acknowledgement

We appreciate the great support by four expert pathologists, Dr. Xiaokang Ke, Dr. Jiacai Ren, Dr. Xiaoyan Wu and Dr. Feng Guan, and the secretary of the Pathology Department, Mrs. Lingli Xia, at Renmin Hospital of Wuhan university. We give special thanks to Dr. Jun Liang in the Department of Nuclear Medicine at Renmin Hospital of Wuhan University and Dr. Jie Tan in the Department of Breast the Thyroid Surgery at Wuhan Union Hospital for their contribution and guidance in patient selection and grouping. We thank Prof. Katherine Hoadley at the University of North Carolina at Chapel Hill for her kind help in building a profile-based scoring system. We thank Dr. Zhou Liu from the Breast Tumor Center of Sun Yat-Sen Memorial Hospital and Dr. Nancy Li from the Reactome HelpDesk for their timely replies on two independent technical problems. We pay special thanks to Prof. Tiannan Guo and Prof. Yi Zhu for providing the platform. We pay special thanks to authors who provided the external expression profiles for validation in the present study.

## Author contribution

H.L. contributed to conceptualization, data curation, formal analysis, investigation, methodology, project administration, visualization and writing - original draft. J.W. contributed to data curation, formal analysis, investigation and writing - original draft. Y.Z. contributed to data curation, formal analysis, methodology, resources, software and writing - original draft. P.H. and L.L. contributed to data curation and software. D.Y. contributed to investigation, validation, visualization and writing – review & editing. D.K. and Z.X. contributed to data curation and investigation. Y.S. contributed to formal analysis, methodology, project administration, resources, software, supervision, writing - original draft and writing – review & editing. C.C. contributed to project administration, resources, supervision, writing – review & editing, conceptualization and funding acquisition.

## Abbreviations

ANOVA: analysis of variance
ATA: American Thyroid Association
AUC: area under the curve (of ROC)
*BRAF*: B-Raf proto-oncogene, serine/threonine kinase
BRS: *BRAF-RAS* score
CC: correlation coefficient
COSMIC: the Catalogue Of Somatic Mutations In Cancer
CT: computed tomography
CYT: immune cytolytic activity score
DEG: differentially expressed gene
DEP: differentially expressed protein
DIA: data-independent acquisition
DTC: differentiated thyroid cancer
ECP: eosinophil cationic protein
ES: enrichment score
ERK: extracellular signal-regulated kinase
ETE: extrathyroidal extension
FC: fold change
FFPE: formalin-fixed paraffin-embedded
F-18-FDG: 2-[^18^F]fluoro-2-deoxy-D-glucose
GEO: the Gene Expression Omnibus database
GO: gene ontology
GS: gene significance
GSEA: gene set enrichment analysis
GSVA: gene set variation analysis
HR: hazard ratio
ID: positive RAI uptake and disease persistence
Id: RAI uptake positive and disease remission
iDF: negative RAI uptake at the first time of RAI treatment with disease persistence
iDG: RAI uptake lost gradually after previous RAI treatments with disease persistence
IIS: immune infiltration score
IP: inositol phosphate
IPA: ingenuine pathway analysis
KEGG: Kyoto Encyclopedia of Genes and Genomes
KNN: K-nearest neighbor
LASSO: least absolute shrinkage and selection operator
LC: liquid chromatography
LNM: lymph node metastatic lesion
MAPK: mitogen-activated protein kinase
MET: MET proto-oncogene
MKI: multi-kinase inhibitor
MM: module membership
MS: mass spectrometry
MRI: magnetic resonance imaging
NIS: sodium iodide symporter
PCA: principal component analysis
PCT: pressure cycling technology
PET: positron emission tomography
RAI: radioactive iodine
RAIR: RAI-refractoriness
RF: random forest
RFS: recurrence-free survival
RMA: robust multiarray average
ROC: receiver operating characteristic
RR-PTC: radioactive iodine refractory papillary thyroid cancer
SPECT: single photon emission computerized tomography
ssGSEA: single sample GSEA
SVM-RFE: support vector machine-recursive feature elimination
TAK1: transforming growth factor-β activated kinase 1
TCGA: The Cancer Genome Atlas Program
*TERT*: telomerase reverse transcriptase
*TERTp*: *TERT* promoter
TDS: thyroid differentiation score
TIS: T cell infiltration score
TNF: tumor necrosis factor
TNGS: targeted next-generation sequencing
TOM: topological overlap matrix
WBS: whole-body scanning
WGCNA: weighted correlation network analysis

