## Supplementary methods, Supplementary figure 1-12, and References for "Molecular and Proteomic Profiles of Radioiodine Refractory Papillary Thyroid Cancer"

[#] Equal contribution.

**Additional discussion**

Four issues warrant additional discussion in supplementary materials.

The first issue is about NIS and dedifferentiation evaluation. Many previous studies have investigated the dedifferentiation mechanism and redifferentiation targets of RR-PTC. NIS is the most important transporter of iodide and is highly expressed in thyroid epithelium (1). Previously, researchers hypothesized that the downregulation of NIS in primary tumors contributed to the loss of RAI uptake in metastatic RR-DTC lesions (2). This hypothesis was soon challenged by the intracellular overexpression of NIS in ~70% of thyroid cancer samples (3). In 2018, Feng *et al.* reported that increased intracellular NIS could exert a non-pump pro-tumorigenic effect (4). The relationship between NIS expression and the RAIR has yet to be determined (5, 6). Owing to the methodological limitation of proteomics, we failed to quantify NIS protein expression. However, seven molecules indicating thyroid differentiation, including PAX8, TG and TPO, were identified. Although the TDS tended to increase in the RR-PTC samples, the difference did not reach statistical significance. Consequently, our results do not fully support that thyroid-specific proteins in primary lesions could be used as diagnostic biomarkers for RR-PTC.

Secondly, most of the molecules identified in the article were not the main regulators of their pathways. However, these three proteins warrant further discussion. There are few reports of S100B in thyroid cancer, but its high expression promotes cancer metastasis via interaction with P53 signaling in other glandular epithelium-derived carcinomas (7). NFKB1 belongs to the well-known NF-κB signaling, which is closely related to cancer initiation and progression. A functional polymorphism in its promoter increases the risk of PTC (8). In addition, FN1 overexpression was found in aggressive thyroid cancer and promoted its migration and invasion (9). The expression of this molecule was greater in *BRAF*-mutated PTCs and was indictive of poor prognosis (10). These proteins are associated with aggressive cancer behaviors and could be promising biomarkers for RR-PTC.

The current study did not focus on the iDG group because of its high intrinsic heterogeneity, but one hypothesis could help explain the process of loss of I-131 uptake in this group. The original tumor may comprise heterogeneous cancer cells with different tolerances to I-131 treatment. I-131 exerted a selective effect on cells, and those with greater tolerance survived, which ultimately results in the formation of negative uptake foci on I-131 WBSs.

With respect to proteomics on RR-DTC, Song et al. performed 2D gel electrophoresis to quantify serum proteins derived from ten PTC patients with lung metastases in 2013 (11). Among > 100 proteins, afamin was significantly downregulated in the serum of RR-PTC patients. Ten years later, another study evaluated protein expression in metastatic lymph node samples from six PTC patients (12). Among the 665 DEPs, CHI3L1 was confirmed to be upregulated in RR-PTC using immunohistochemistry. Subsequent cell experiments suggested that CHI3L1 overexpression could activate the classic MAPK pathway, which may then cause NIS dysfunction. These two precursor studies provided novel insights but were limited by small sample sizes and the lack of primary tumors.

**Supplementary methods**

1 Proteomic data acquisition and preprocessing

1.1 Batch design

The quantitative analysis of protein profiles in 168 samples was estimated to take more than one week. For minimization of the potential batch effect due to slight uncontrolled changes in instrument conditions, samples with tumors were allocated into 10 batches using Proteome Expert webserver (13). The first nine batches contained eighteen samples, whereas the remaining six samples were allocated to the tenth batch. In each batch, one sample was randomly selected as the technical replicate to test the intra-batch stability of the sample preparation and data acquisition instruments. In addition, a homogenous pooled thyroid sample, which contained mixed thyroid tissue peptides, was distributed equally to each batch to test the inter-batch stability of the mass spectrometry (MS).

1.2 Protein extraction and digestion

The FFPE samples were prepared for subsequent proteomic analysis as described previously (14-16). Tumors on tissue slides were carefully scraped off and then dewaxed and rehydrated in heptane and ethanol series (100%, 90% and 75%) at room temperature. Next, protein decrosslinking was processed with tris-HCl (100 mM, pH = 10.0) at 95°C for 30 mins and subsequently cooled to 4°C.

Samples were lysed by the mix buffer containing urea, thiourea, tris (2-carboxyethyl) phosphine and iodoacetamide in 100 mM triethylammonium bicarbonate using pressure cycling technology (PCT) (45000 psi, 90 cycles of 30 s high pressure and 10 s ambient pressure, 30°C) in Barocycler (PressureBioSciences Inc., USA). Protein digestion was performed through Lys-C (enzyme-to-substrate ratio: 1:100, Hualishi Tech. Ltd., China) and trypsin (enzyme-to-substrate ratio: 1:50, Hualishi Tech. Ltd., China) in the PCT instrument (20000 psi, 120 cycles of 50 s high pressure and 10 s ambient pressure, 30°C). Trifluoroacetic acid (1%, Thermo Fisher Scientific, USA) was then added to terminate the digestion. The peptide solutions were desalted using SOLAμ solid-phase extraction plates (Thermo Fisher Scientific, USA) following the manufacturer's instructions. NanoScan (Analytic Jena, Germany) was used to measure peptide concentrations at $A_{280}$. Peptide solutions were stored at 4°C for subsequent analysis. All the chemical reagents mentioned above, unless specified, were from Sigma-Aldrich, USA.

1.3 Proteomic data acquisition and library search

Next, 200 ng peptides were injected into a liquid chromatography (LC) system coupled with a trapped ion mobility spectrometry mass spectrometer (Bruker Daltonics, Bremen, Germany) for each run. Prepared peptides were first loaded on to a 15 cm × 75 μm C18 column (1.9 μm, 100 Å, 20 mm × 75 μm i.d.) and the LC effective gradient was 60 mins at a flowrate of 300 nL/min. The linear gradient was composed by buffer B (%) from 5 to 27% and followed by 27-40% within 10 mins and a further boost to 80%. The ion mobility underwent scanning within the range of 0.7 to 1.3 Vs/cm$^2$. MS1 and MS2 acquisition was performed in the range of *m/z* from 100 to 1700 Th. All the chemical reagents and instruments mentioned above, unless specified, were obtained from Thermo Fisher Scientific, USA.

A total of 187 effective data-independent acquisition (DIA) files including 9 technical replicates and 10 pooled samples were acquired. These raw files were then analyzed against a thyroid tissue specific spectral library (15) using DIA-NN (v1.8.1) (17). Fixed and variable modifications were set as cysteine carbamidomethylation and methionine oxidation, respectively. Peptide length range, precursor *m/z* range and fragment ion *m/z* range were set as 7–30, 300–1800, and 200–1800, respectively. False discovery rate of the precursor was set to 1%. Other parameters remained as default values.

1.4 Quality control and data preprocessing

Principal component analysis (PCA) was conducted using the *stats* R package (v3.6.0) to visualize the batch effect of proteomic profiles. Pearson correlation coefficients were calculated using the expression profiles of the technical replicates and their original samples. Similarly, correlation analysis was conducted to assess the batch effect using homogenous pooled thyroid sample tissues. Next, proteins with missing values less than 50% were regarded as quantifiable and forwarded for data normalization. The K-nearest neighbor (KNN) algorithm was applied for the missing value imputation using the *SeqKnn* R package. Since batch effects were undetectable, data correction was not performed. UniProtKB AC/IDs were converted to official gene symbols using the UniProt database (v2023.03) (18). When multiple IDs corresponded to one gene symbol, the median was calculated. Tumor purity was analyzed by the *ESTIMATE* algorithm.

2 Targeted next-generation sequencing (TNGS)

The primary lesions from 48 patients with PTC were subjected to molecular profiling via TNGS. The protocol was performed as described previously (19). Briefly, DNA was extracted from FFPE tissue sections using the QIAamp DNA FFPE Tissue Kit (Qiagen, Germany) following the manufacturer's instructions. After amplification of targeted DNA fragments and removal of primers, the products were purified using an Ion AmpliSeq Library Kit (Thermo Fisher Scientific, USA). The concentrations of samples were quantitated by a NanoDrop system (Thermo Fisher Scientific, USA). The quality of the purified DNA was evaluated by 1% agarose gel electrophoresis. Severe degradation was detected in three samples, which were excluded from further analysis.

The remaining 45 library products were sequenced via 150 bp paired-end runs on the NextSeq 500 platform (Illumina, Inc., USA). The medians of sequencing depth and coverage were 5136× and 98.7%, respectively. Sequencing data were aligned to a reference human genome dataset (hg19/GRCh37). Subsequently, read mapping, quality control, variant calling and genotyping were performed following the protocols of the OncoAim® Thyroid Cancer Multigene Assay Kit (Singlera Genomics, Inc., China). Mutations and fusions were evaluated for 23 genes (Sup table S1). The minimum confidence threshold for variant allele frequency was 5%. The ENSEMBL Variant Effect Predictor (v90) was used for variant functional annotation. In addition, the gene variants were searched against the ClinVar database (v2020006) (20). Somatic gene variants were categorized for clinical significance based on the Catalogue Of Somatic Mutations In Cancer database (COSMIC, v97) (21). Of note, gene variants with unknown clinical significance were subjected to nonparametric tests after those with a frequency < 3 were filtered out.

### 3 Bioinformatic analysis

#### 3.1 Dimension reduction and differential expression analysis

As depicted above, the raw data were converted to a normalized expression profile. The dimension of the profile was reduced and visualized using the principal component analysis (PCA) algorithm with the *stats* R package (v3.6.0).

Differential expression analysis was conducted using the *limma* R package (v3.56.2). Proteins with absolute fold change (FC) > 1.414 and *P* value < 0.05 were defined as differentially expressed proteins (DEPs). Heatmaps were drawn using the *pheatmap* R package (v1.0.12). Samples were clustered with the Euclidean distance and complete method.

Analysis of variance was used to identify proteins correlated with severity of RR-PTC. Proteins with statistical significance were clustered using the *mfuzz* R package (v3.19). Proteins in monotonic regulated clusters were regarded as DEPs.

#### 3.2 Gene ontology (GO) and pathway enrichment analysis

Gene set enrichment analysis (GSEA) was conducted using GSEA software (v4.3.2) to evaluate potential pathways and molecular mechanisms (22). Expression profiles and phenotype labels were loaded into the software, and annotated gene sets, such as Reactome and Kyoto Encyclopedia of Genes and Genomes (KEGG) annotation files, were downloaded from the Molecular Signatures Databases (23). The minimum and maximum for excluding gene sets were set as 5 and 5000, respectively. The other parameters were set to their defaults. A *P* value < 0.05 was considered to indicate a significant difference. The enrichment scores (ESs) were normalized to the mean enrichment of random samples of the same size.

For gene set variation analysis (GSVA), ESs were calculated with the *GSEA* R package (version 1.40.1) (24). Predefined gene sets were downloaded as described above and the settings used were the same as those used for GSEA. ES profiles were produced and then evaluated for differential expression using the *limma* package. A *P* value < 0.05 was considered the threshold for a significant difference.

The top GO processes were enriched by Metascape web-based platform (25). Annotations were set as default. Ingenuine pathway analysis (IPA) of the regulated proteins identifies most significantly relevant pathways with *P* value of determined based on right-tailed Fisher's Exact Test with the overall activation or inhibition states of enriched pathways were predicted by *z*-score (26).

#### 3.3 Immune infiltration analysis

The proportions of 22 immune cells types were calculated using the *CIBERSORTx* algorithm (27). For evaluation of the purity of the tumor tissues, estimated scores were calculated for each sample with the *ESTIMATE* R package (v2.0.0) (28). Immune cytolytic activity score (CYT) was calculated from geometric means of normalized expression levels of the two effector genes (granzyme A and perforin 1) (29).

In addition, the *single sample GSEA* (*ssGSEA*) algorithm in the *GSVA* R package (v 3.19) (30) was used to estimate the proportions of 28 immune cell types (31). T cell infiltration score (TIS) and immune infiltration score (IIS) were calculated using the *Z*-score-standardized *ssGSEA* matrix, as depicted in a previous study (32).

#### 3.4 Evaluation of tumor differentiation and extracellular signal-regulated kinase (ERK) pathway activity

The thyroid differentiation score (TDS) was developed based on sixteen genes that closely related to thyroid function (33). Seven genes were identified in our profile (DUOX1, DUOX2, FOXE1, NKX2-1, PAX8, TG and TPO). The log2-normalized values were first centered at the median across samples. The means across seven genes were then calculated for each sample. Since these genes either encode thyroid-specific proteins or are highly expressed in thyroid follicular cells, lower TDSs indicate poor differentiation of thyroid cancer tissues.

The extracellular signal-regulated kinase (ERK) pathway is essential for the uncontrolled proliferation of *BRAF*-mutated tumor cells. The ERK score was calculated to evaluate the activity of the ERK pathway (34). A total of eighteen genes involved in the scoring system were identified in the current analysis. The mean Z scores of the eighteen genes were computed and a higher score indicated more activated pathways (32).

#### 3.5 Weighted correlation network analysis (WGCNA)

WGCNA is an analytical tool for describing correlation patterns among genes across large-scale, high-dimensional datasets. Genes with high correlation were clustered to form a module. The modules were then related to external sample traits using the eigengene network methodology. The subsequent analysis was conducted using the *WGCNA* R package (v1.72-1) (35). Since genes were filtered during data preprocessing, all 7394 genes were processed for network construction to retain as much information as possible. No outlier sample was identified using the goodSamplesGenes function (Sup figure 10A). A correlation matrix was constructed based on pairwise genes with Pearson's correlation coefficient and average linkage method. The matrix was then weighted using a power function to form an adjacency matrix. The soft threshold β was determined to be 3 by calculating the scale independence and mean connectivity to emphasize strong correlations between genes and penalize weak correlations (Sup figure 10B&C). The adjacency matrix was then converted to a topological overlap matrix (TOM). A gene dendrogram was constructed using average linkage hierarchical clustering to classify genes with similar expression profiles (Sup figure 10D). A total of 21 significant modules were identified at minimum module size = 30, merge cut height = 0.25 and deepSplit = 3 using the cutreeDynamic function. The expression of each module was summarized by the module eigengene (Sup figure 10E). Gene significance (GS) was defined as the correlation between the gene and the trait of interest, while module membership (MM) represented the correlation between the module eigengenes and the gene expression profile. Sample mutation traits were subsequently input for the subsequent assessment of GS and MM (Sup figure 10F).

#### 3.6 Machine learning algorithms

Least absolute shrinkage and selection operator (LASSO) regression, random forest (RF), and support vector machine-recursive feature elimination (SVM-RFE) were used to identify biomarker proteins for gene mutations and fusions (36-38).

LASSO was conducted using the *glmnet* R package (v4.1-8). After model construction, lambda values were set at the minimum mean-squared error to select prominent variables (Sup figure 11A).

RF was conducted using the *caret* R package (v6.0-94) with fivefold cross-validation. The correlation between the accuracy of repeated cross validation along with the number of genes was calculated for gene selection (Sup figure 11B).

SVM-RFE was conducted using the *e1071* R package (v1.7-13) with fivefold cross validation. The relation of error against gene number was plotted for gene selection (Sup figure 11C).

#### 3.7 External dataset validation

Several external datasets were used for the validation of our results. GSE151179, which was published on May 26, 2020, by Colombo and colleagues (39), included 52 samples derived from radioiodine-refractory and radioiodine-avid PTC patients. RNA was extracted from snap-frozen tissues and then quantified with a gene/miRNA microarray. The sample types included primary lesions, positive lymph nodes and matched normal tissues. The raw data were downloaded from the Gene Expression Omnibus (GEO) database (40) and preprocessed according to the authors' instructions. Medians were calculated when multiple transcripts corresponded to one gene. Primary samples were pooled and labeled according to their I-131 uptake ability and response to radiotherapy. GSVA was conducted as described in the previous section. The log-transformed values were subjected to the Mann-Whitney *U* test for gene expression.

The Cancer Genome Atlas (TCGA) published information on the genomic landscape, gene expression and other molecular features of 496 PTCs in 2014 (33). The patient number was increased to 504 at the time of data extraction. The transcript profiles of 571 samples were downloaded from the TCGA-THCA program. RNA-seq raw counts were extracted from the raw data files and then merged into one expression matrix using the *tidyverse*, *vroom* and *jsonlite* R packages. A total of 22,418 of 60,660 transcripts with a missing ratio > 0.8 were excluded from subsequent analyses. The data were normalized with the trimmed-mean of M values method in *edgeR* package (v4.0.2) and transformed to log2+1 counts per million mapped reads. Primary lesion samples were included for subsequent analysis.

TCGA samples were labeled with gene variances. Single nucleotide variants and insertions and deletions identified using whole exome DNA sequencing were collected from TCGA. The tiers of gene mutations were determined based on the COSMIC database as described above (21). Information on gene fusions was collected from the ChimerDB 4.0 database (41). This database included all samples in TCGA, and gene fusion was assessed by analyzing deep sequencing data.

Raw data from four external datasets were downloaded from the GEO database (Sup Table S7) (42-44) and normalized using the robust multiarray average (RMA) method. The PTC samples with *BRAF*, *RAS* and/or fusion status data were retained for data preprocessing. Batch effects among the four matrices were removed using the *sva* R package (v3.20.0) (Sup figure 12). Medians were calculated when multiple transcripts corresponded to one gene. Differential expression analyses were conducted for TCGA- and multiple GEO-derived expression profiles. The threshold of the adjusted *P* value was set as 0.05.

The predictive performance of the selected genes in the external datasets was evaluated with receiver operating characteristic (ROC) curves. The area under the curve (AUC) was calculated using the *pROC* R package (v1.18.5).

#### 3.8 Survival analysis

Survival analysis was conducted using GEPIA2, an online tool based on the TCGA database (45). The THCA project was selected. Since most PTCs do not lead to death even with relapse or metastasis, recurrence-free survival (RFS) was evaluated instead of overall survival. The hazard ratio (HR) and *P* value were calculated.

### 4 Immunohistochemistry

FFPE tissue sections (5 μm) were prepared for immunohistochemistry (if sample quantity permitted) with the following antibodies: anti-ECP (eosinophil cationic protein; GB112150), anti-CD11c (GB115690), anti-CD27 (GB111165) and anti-CD68 (GB113109). The four antigens were identified as biomarkers for eosinophils, immature dendritic cells, memory B cells and macrophages, respectively (46-49). The sections were stained using a DAB substrate kit (G1212) according to the manufacturer's instructions. An expert pathologist, blinded to the sample groups, evaluated each IHC section. The number of positive immune cells was counted manually in five randomly selected fields at ×200 magnification, and the average was then calculated. All the antibodies and chemical reagents were sourced from Servicebio Co., Ltd., China.

**Supplementary tables**

Supplementary tables. S1-S7

Details are provided in a Microsoft excel file.

**Supplementary figures**

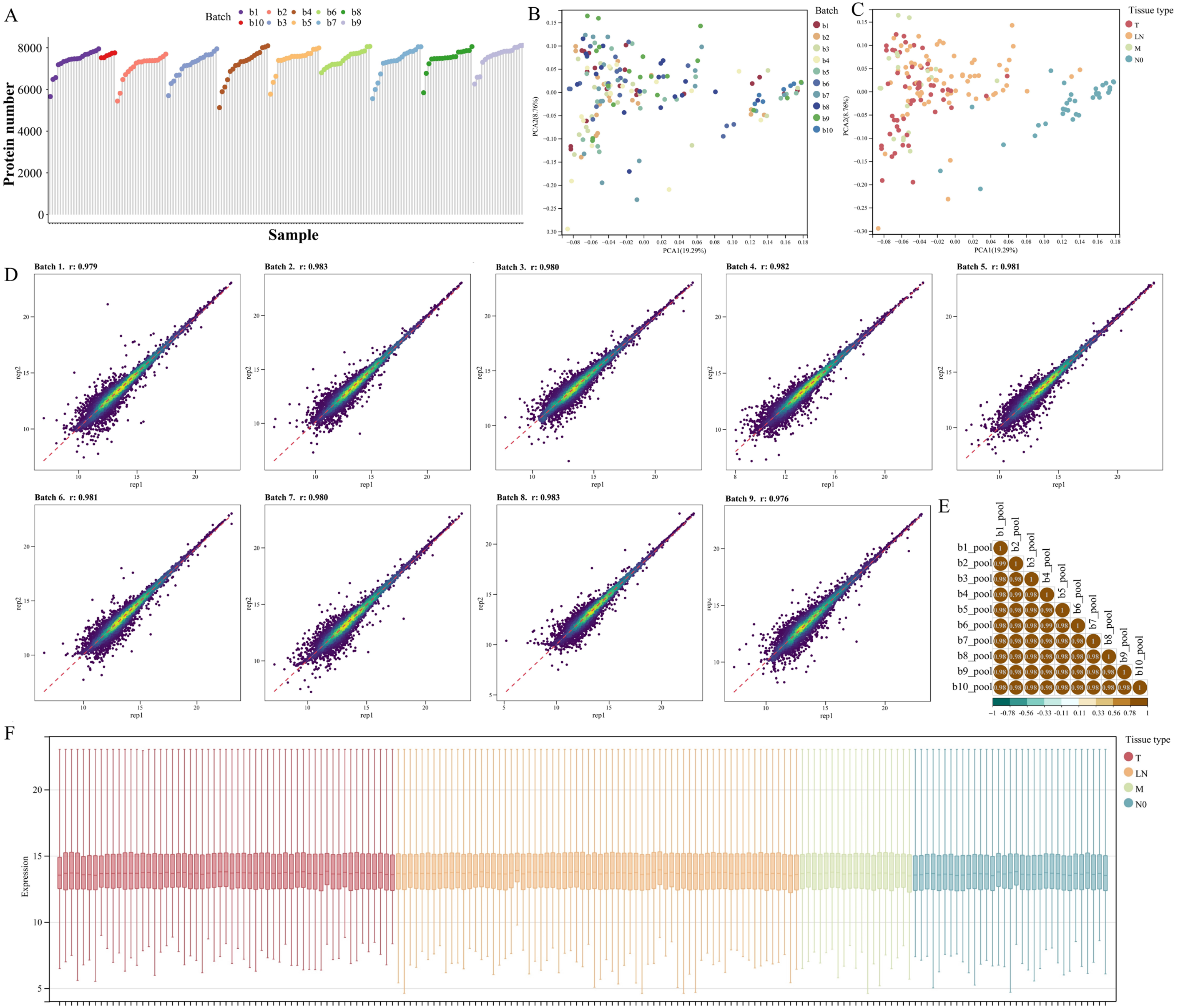
A
Batch
b1
b2
b3
b4
b5
b6
b7
b8
b9
b10
Protein number
Sample
B
Batch
PCA1(19.29%)
PCA2(8.76%)
C
Tissue type
T
LN
M
N0
PCA1(19.29%)
PCA2(8.76%)
D
Batch 1. r: 0.979
Batch 2. r: 0.983
Batch 3. r: 0.980
Batch 4. r: 0.982
Batch 5. r: 0.981
Batch 6. r: 0.981
Batch 7. r: 0.980
Batch 8. r: 0.983
Batch 9. r: 0.976
rep1
rep2
E
b1_pool
b2_pool
b3_pool
b4_pool
b5_pool
b6_pool
b7_pool
b8_pool
b9_pool
b10_pool
F
Tissue type
T
LN
M
N0
Expression

Supplementary figure 1. The quality control of proteomic analysis. (A) The number of proteins detected in each sample. A total of 9769 proteins were identified, which indicated the high quality of our data. The minimum and maximum protein numbers were 5120 (52.4%) and 8106 (83.0%), respectively, with a mean of 7481 (76.6%). Since the number of proteins identified in all the samples exceeded the threefold interquartile range (1614), no sample was excluded from subsequent analysis. (B&C) PCA diagram of analyzed samples labeled with batch and tissue type. (D) Pearson correlation analysis in each batch using technical replicates. (E) Pearson correlation analysis of batch effects using pooled thyroid samples. (F) The normalized expression of protein profiles labeled with tissue types. No observable batch effect was detected.

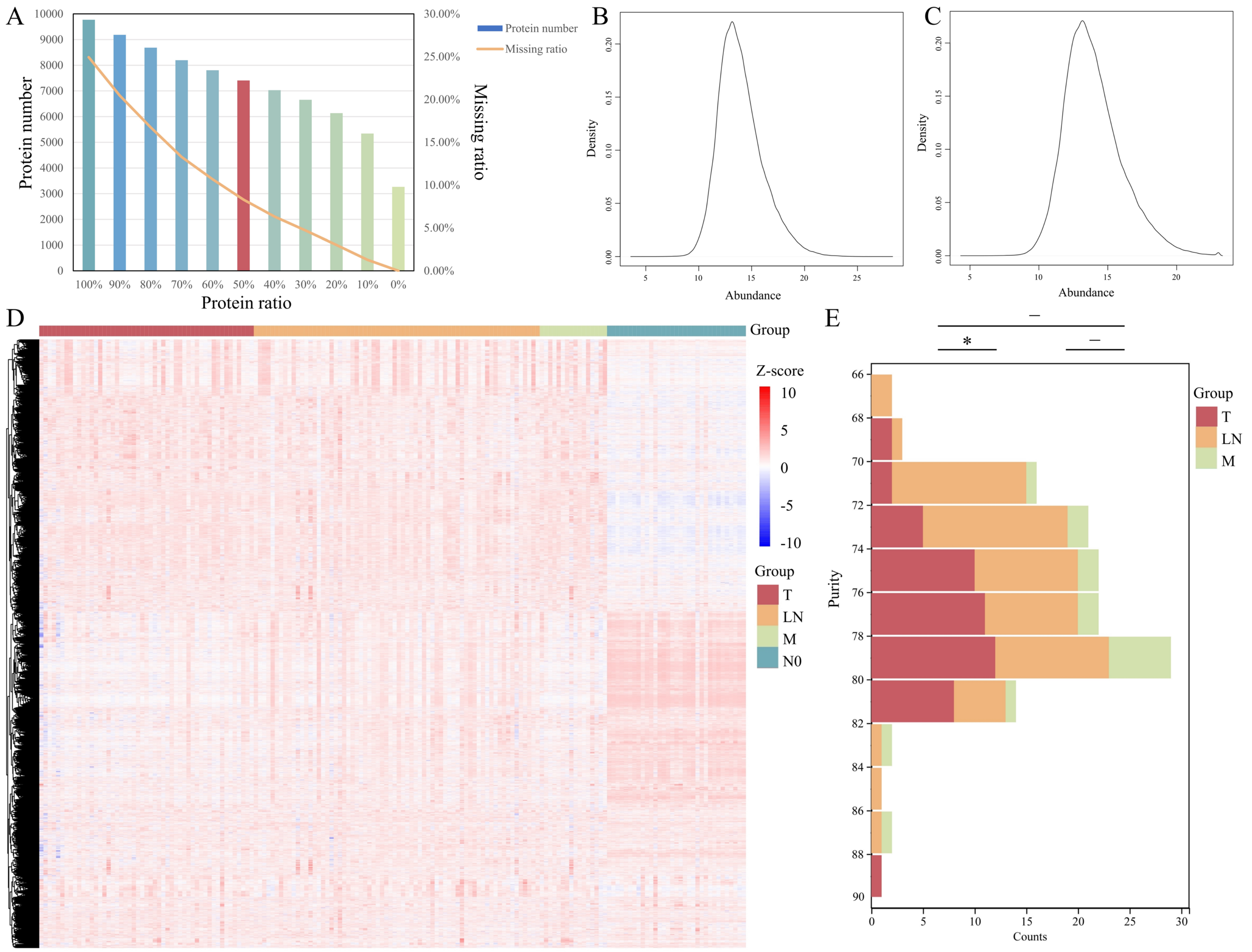

A
Protein number
Missing ratio
Protein ratio
Protein number
Missing ratio
100% 90% 80% 70% 60% 50% 40% 30% 20% 10% 0%
B
Density
Abundance
C
Density
Abundance
D
Group
Z-score
10
5
0
-5
-10
Group
T
LN
M
N0
E
*
Purity
Counts
Group
T
LN
M

Supplementary figure 2. Preprocessing of proteomic profiles. (A) The protein numbers and their corresponding missing ratios. The percentage of missing proteins in the expression matrix decreased from 24.94% to 8.30% after 2362 proteins with a missing ratio > 50% were excluded (red bar). Missing values were not found for 3265 proteins. (B&C) Density plots of protein abundance before and after the removal of outliers. A total of 1079 (0.0823%) of outliers in the whole matrix were substituted. (D) The full expression profiles displayed 168 samples and 7394 proteins with gene symbols after data preprocessing. The mean, median, median, lower and higher quartiles of the log-transformed matrix were 14.4580, 14.1805, 13.0386 and 15.6017, respectively. (E) The tumor purity of samples with cancer tissues calculated using the *ESTIMATE* algorithm. The Kruskal-Wallis test was applied to analyze the distribution of tumor purity across the three groups.

* The distribution of purity in primary tumors and cervical LNMs significantly differed (adj. $P = 0.038$).

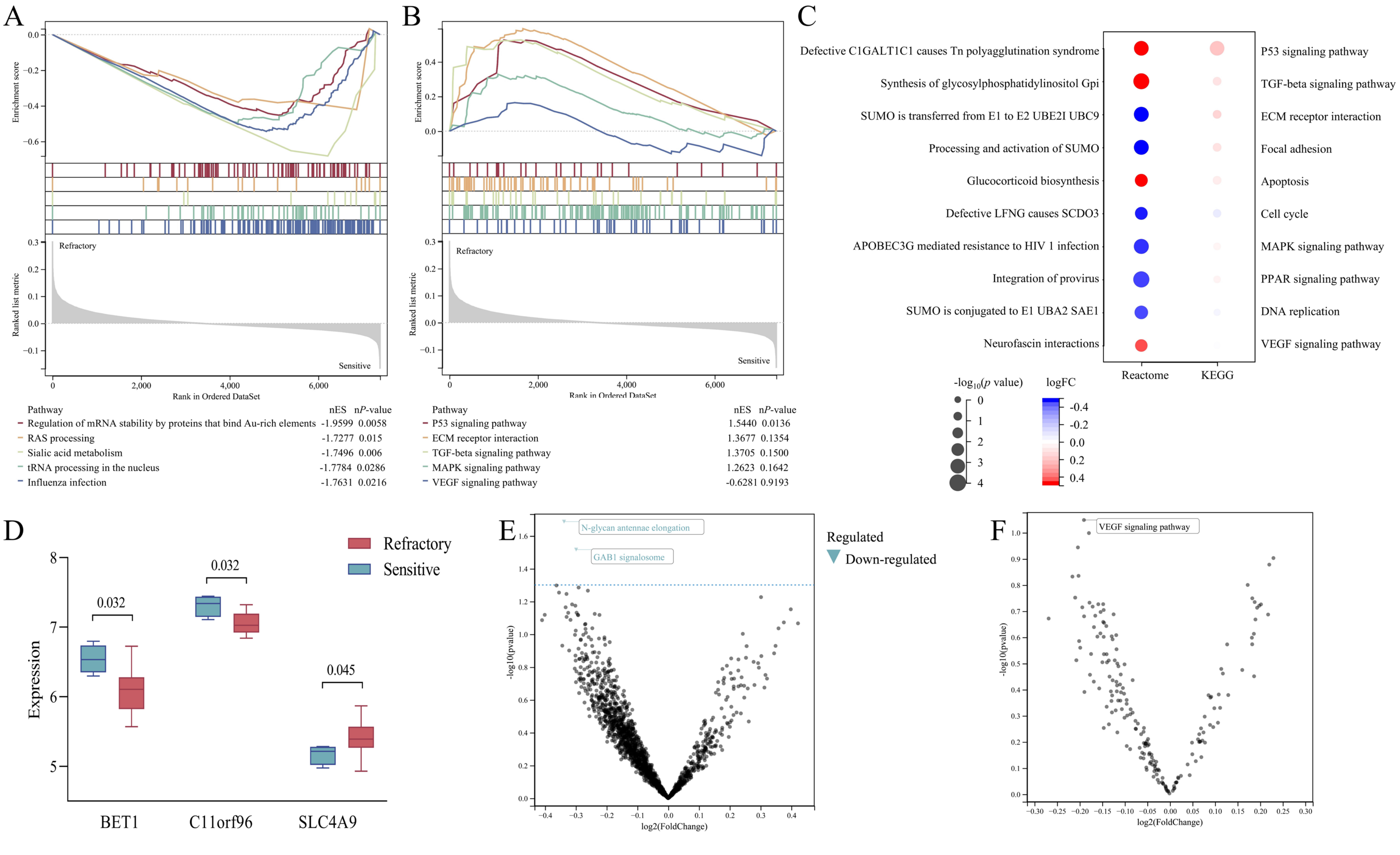
A
Enrichment score
Ranked list metric
Refractory
Sensitive
Rank in Ordered DataSet
Pathway nES nP-value
Regulation of mRNA stability by proteins that bind Au-rich elements -1.9599 0.0058
RAS processing -1.7277 0.015
Sialic acid metabolism -1.7496 0.006
tRNA processing in the nucleus -1.7784 0.0286
Influenza infection -1.7631 0.0216
B
Enrichment score
Ranked list metric
Refractory
Sensitive
Rank in Ordered DataSet
Pathway nES nP-value
P53 signaling pathway 1.5440 0.0136
ECM receptor interaction 1.3677 0.1354
TGF-beta signaling pathway 1.3705 0.1500
MAPK signaling pathway 1.2623 0.1642
VEGF signaling pathway -0.6281 0.9193
C
Defective C1GALT1C1 causes Tn polyagglutination syndrome
Synthesis of glycosylphosphatidylinositol Gpi
SUMO is transferred from E1 to E2 UBE2I UBC9
Processing and activation of SUMO
Glucocorticoid biosynthesis
Defective LFNG causes SCDO3
APOBEC3G mediated resistance to HIV 1 infection
Integration of provirus
SUMO is conjugated to E1 UBA2 SAE1
Neurofascin interactions
P53 signaling pathway
TGF-beta signaling pathway
ECM receptor interaction
Focal adhesion
Apoptosis
Cell cycle
MAPK signaling pathway
PPAR signaling pathway
DNA replication
VEGF signaling pathway
Reactome
KEGG
-log10(p value)
logFC
D
Refractory
Sensitive
Expression
BET1
C11orf96
SLC4A9
0.032
0.032
0.045
E
N-glycan antennae elongation
GAB1 signalosome
Regulated
Down-regulated
-log10(pvalue)
log2(FoldChange)
F
VEGF signaling pathway
-log10(pvalue)
log2(FoldChange)

Supplementary figure 3. Pathway enrichment analysis and validation. (A) The five most enriched pathways identified by GSEA using the Reactome annotation file. (B) Five pathways related to the oncogenesis of the thyroid follicular epithelium. (C) Dot plot based on GSVA. The top ten enriched pathways and those related to oncogenesis and dedifferentiation are displayed in left and right columns, respectively. (D) Three DEGs/DEPs were differentially expressed in both GSE151179 and the present study. Ninety-nine of the 107 DEPs were identified in this cohort. BET1 median: 6.534 *vs.* 6.105, $P = 0.032$; C11orf96 median: 7.338 *vs.* 7.028, $P = 0.032$; SLC4A9 median: 5.216 *vs.* 5.392, $P = 0.045$. (E&F) Volcano maps of GSVA based on the Reactome and KEGG databases, respectively. Two pathways, N-glycan antennae elongation and GAB1 signalosome, were dysregulated in the external transcriptomic profile but not in the current proteomic study.

Abbreviations: nES, normalized enrichment score; n*P*-value, normalized *P* value.

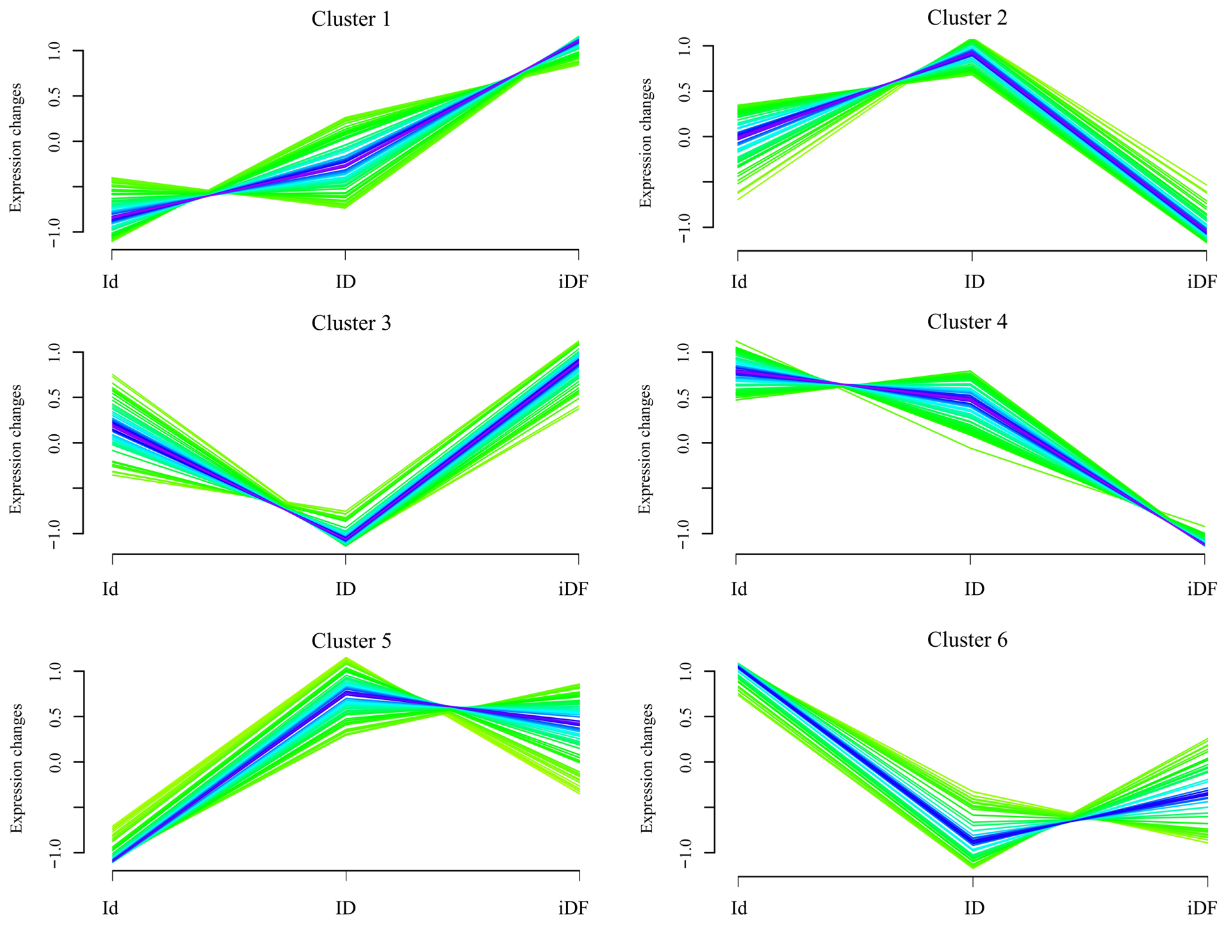

Supplementary figure 4. The clustering of 403 proteins along with the severity of RR-PTC. Proteins with significance difference in analysis of variance were divided into six clusters Proteins in cluster 1 and 4 were monotonic regulated.

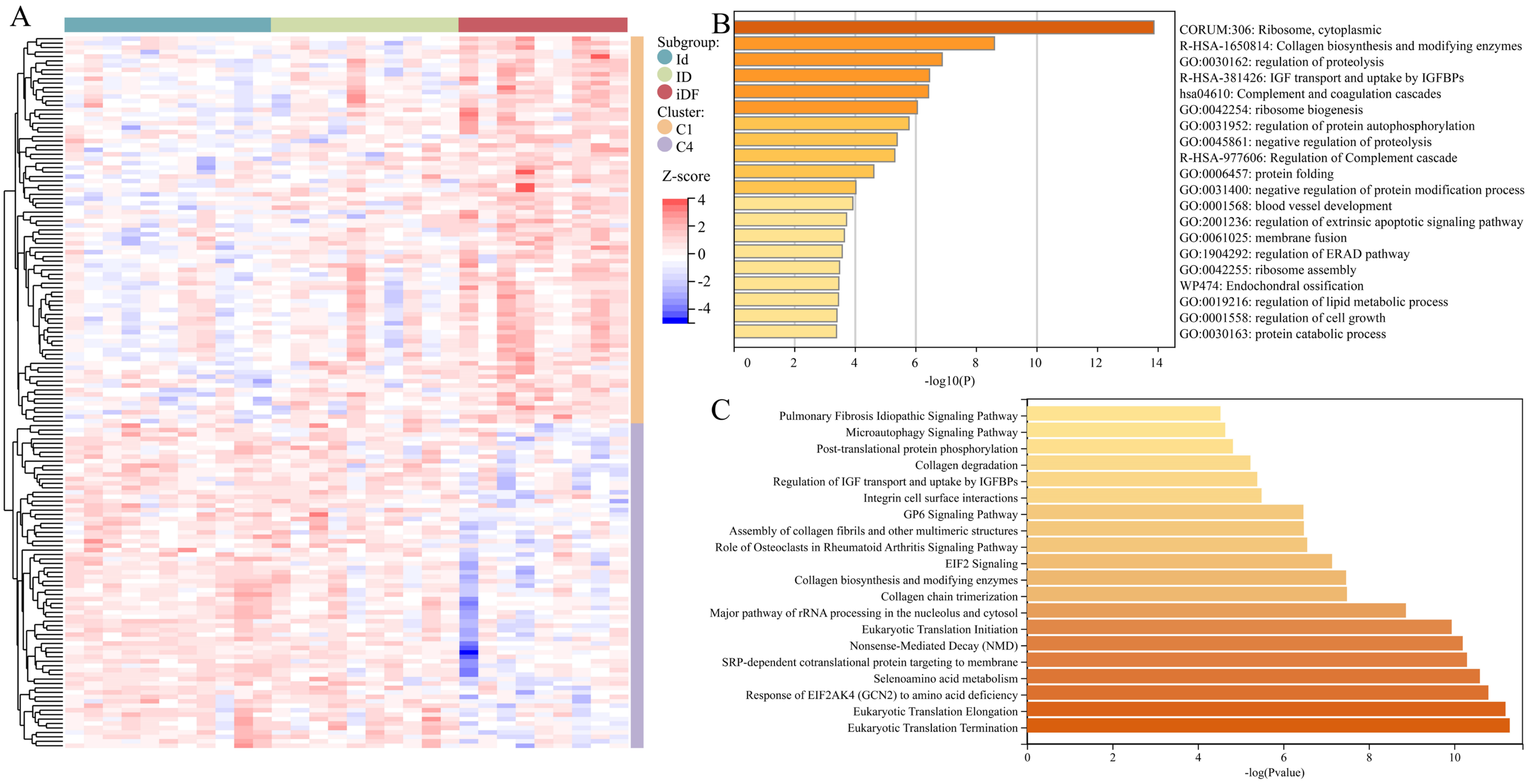

Supplementary figure 5. The gene ontology and pathway enrichment analysis of monotonic regulated proteins. (A) The expression of 160 proteins in Id, ID and iDF subgroups. (B) The top non-redundant gene ontology enriched by Metascape. (C) IPA of top 20 most significantly relevant pathways.

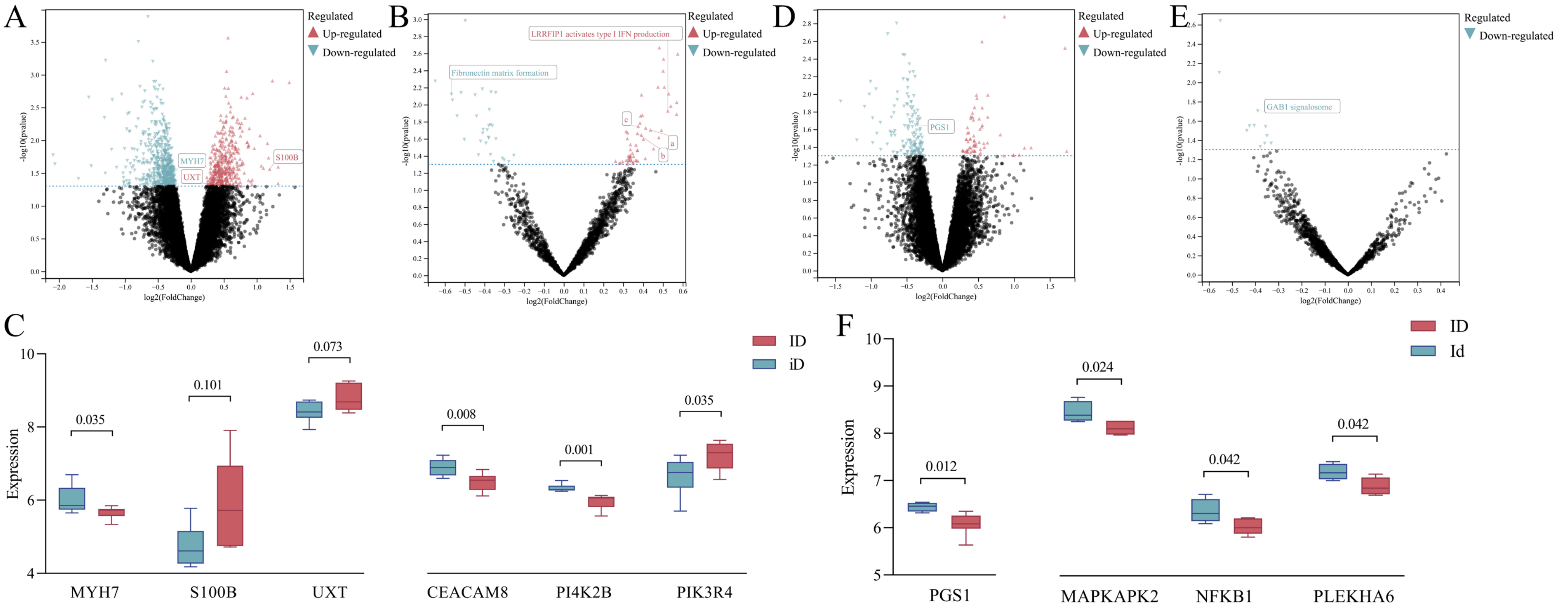

Supplementary figure 6. Validation of DEPs and selected pathways. (A) Volcano map of genes in comparison of the ID and iD groups. Three genes were differentially expressed in both our proteomic profiles and external transcriptomic profiles. However, the regulatory direction was completely different. In the proteomic profile, MYH7 was upregulated, whereas in the transcriptomic profile, its expression was greater in the iD group. S100B and UXT also exhibited inverse changes in the two profiles. (B) Volcano map of pathways identified by comparing the ID and iD groups. Five Reactome pathways were enriched in the two profiles. a, IRAK2 mediated activation of TAK1 complex; b, TICAM1, TRAF6-dependent induction of TAK1 complex; c, TRAF6-mediated induction of TAK1 complex within TLR4 complex. (C) The log-transformed expression of six genes in the transcriptomic profile. MYH7, among all common DEGs, significantly differed according to the nonparametric test ($P = 0.035$). For proteins in selected and common pathways, PI4K2B and PIK3R4 participate in the synthesis of PIPs at the early endosome membrane, and CEACAM8 is involved in fibronectin matrix formation. (D) Volcano map of genes in the comparison of ID and Id groups. PGS1 was differentially expressed between the two profiles (median: 6.080 *vs.* 6.456, $P = 0.012$). (E) Volcano map of pathways identified by comparing the ID and Id groups. GAB1 signalosome was enriched in two profiles ($P = 0.012$). (F) The log-transformed expression of four genes in the transcriptomic profile. Among those involved in the above thirteen pathways, three genes (MAPKAPK2, NFKB1 and PLKHA6) were downregulated in the ID group, in contrast to the expression pattern observed in our proteomic profile.

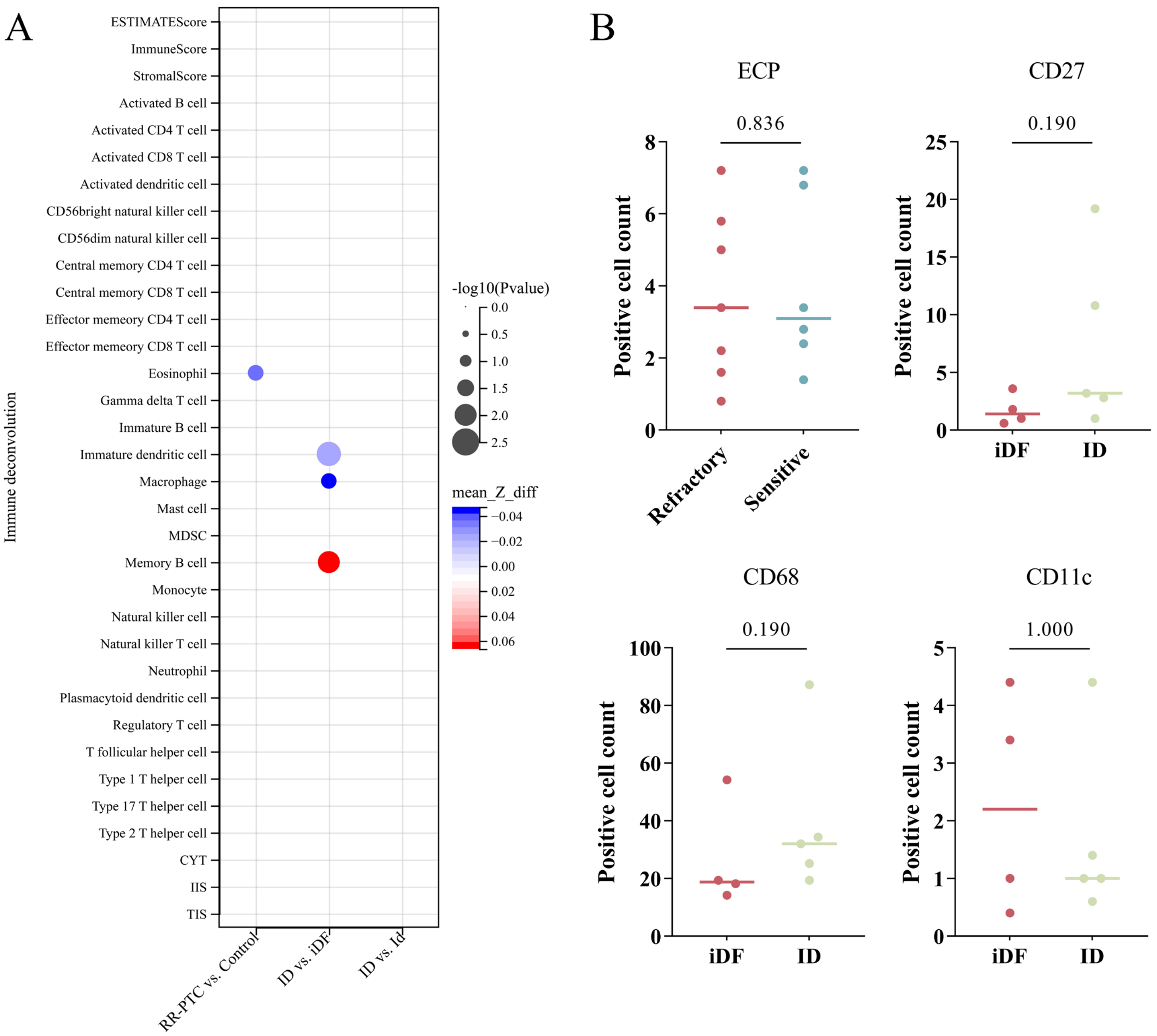

Supplementary figure 7. Tumor microenvironment feature discriminating RR-PTC *vs.* Control, ID *vs.* IDF and ID *vs.* Id. Immune analysis was conducted based on the ssGSEA matrix. (A) Memory B cell, macrophage and immature T cell were related to I-131 uptake. The comparison was based on the ssGSEA matrix. (B) Immunohistochemistry did not show any statistical significance on immune cells.

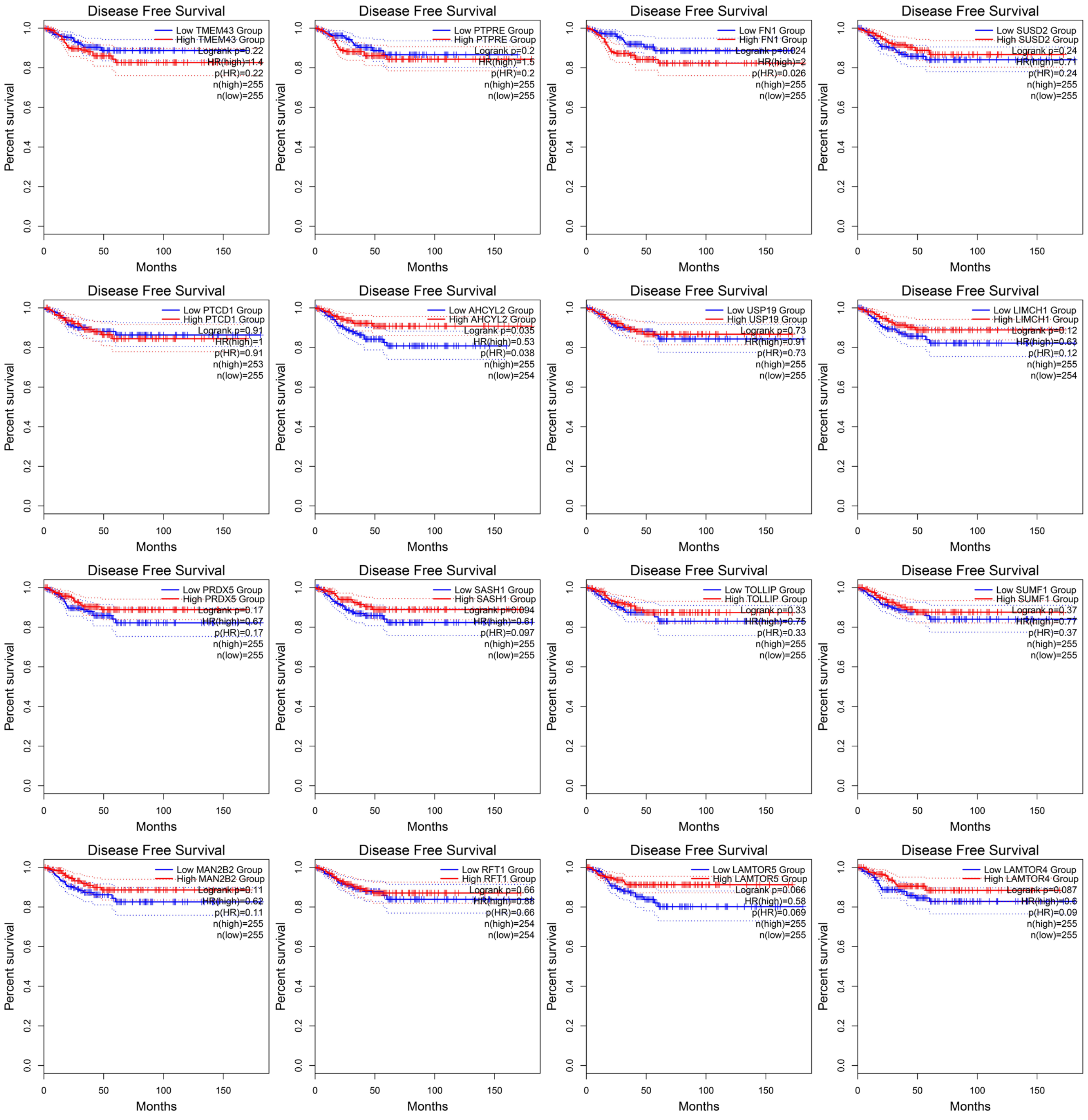

Supplementary figure 8. Survival analysis of selected biomarkers. RFS was defined as the time interval between the date of diagnosis and the date of relapse/recurrence or death (all causes) whichever occurred first.

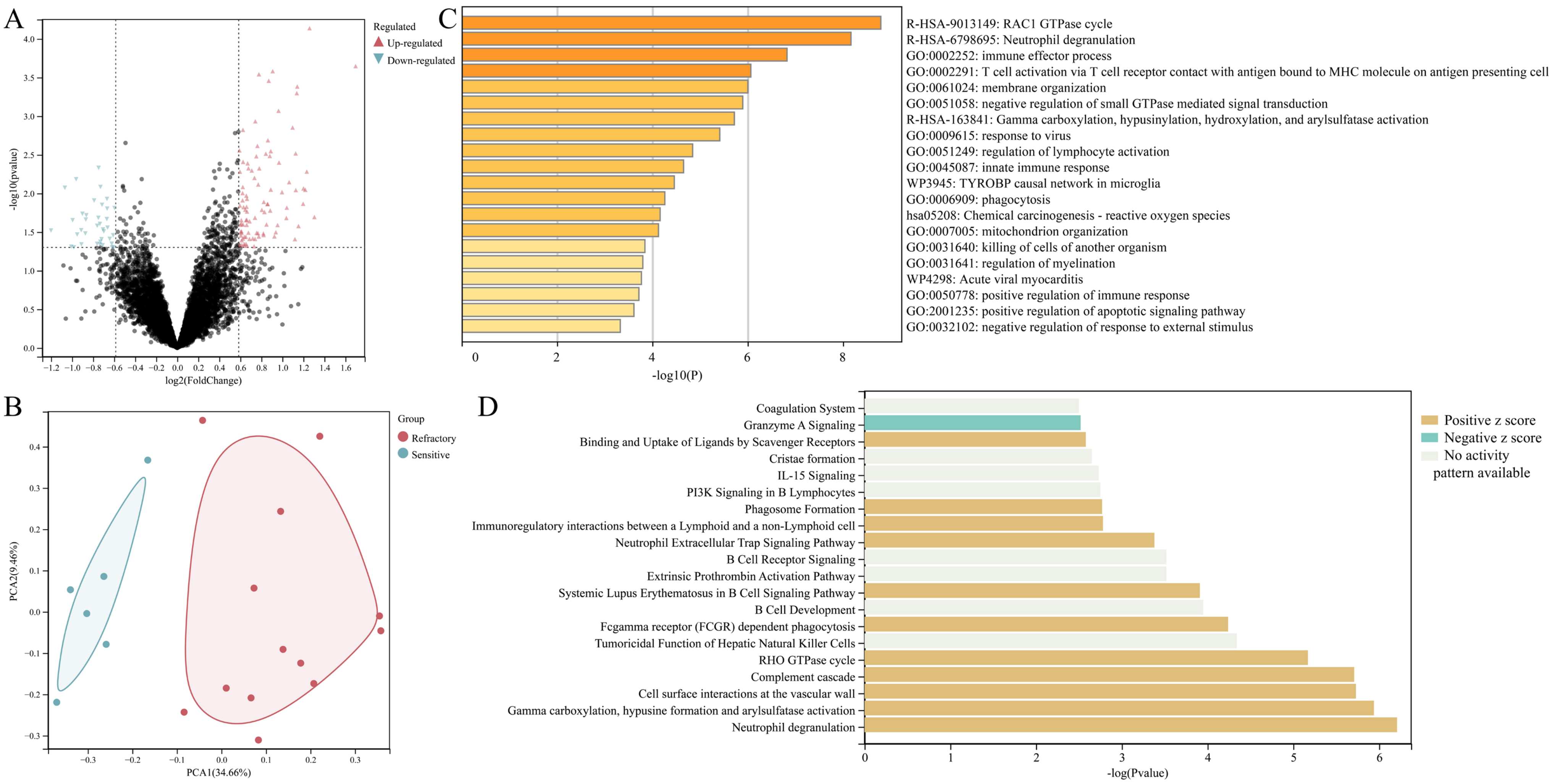

Supplementary figure 9. The differential expression analysis, gene ontology and pathway enrichment analysis for samples with *BRAF* mutation and *TERTp* wild type. The RR-PTC and control groups were compared. (A) The differential expression analysis identified 184 DEPs. (B) Dimension reduction visualization of two groups. (C) The top non-redundant gene ontology enriched by Metascape. (D) Ingenuine pathway analysis of most significantly relevant pathways with the predicted activation or inhibition state.

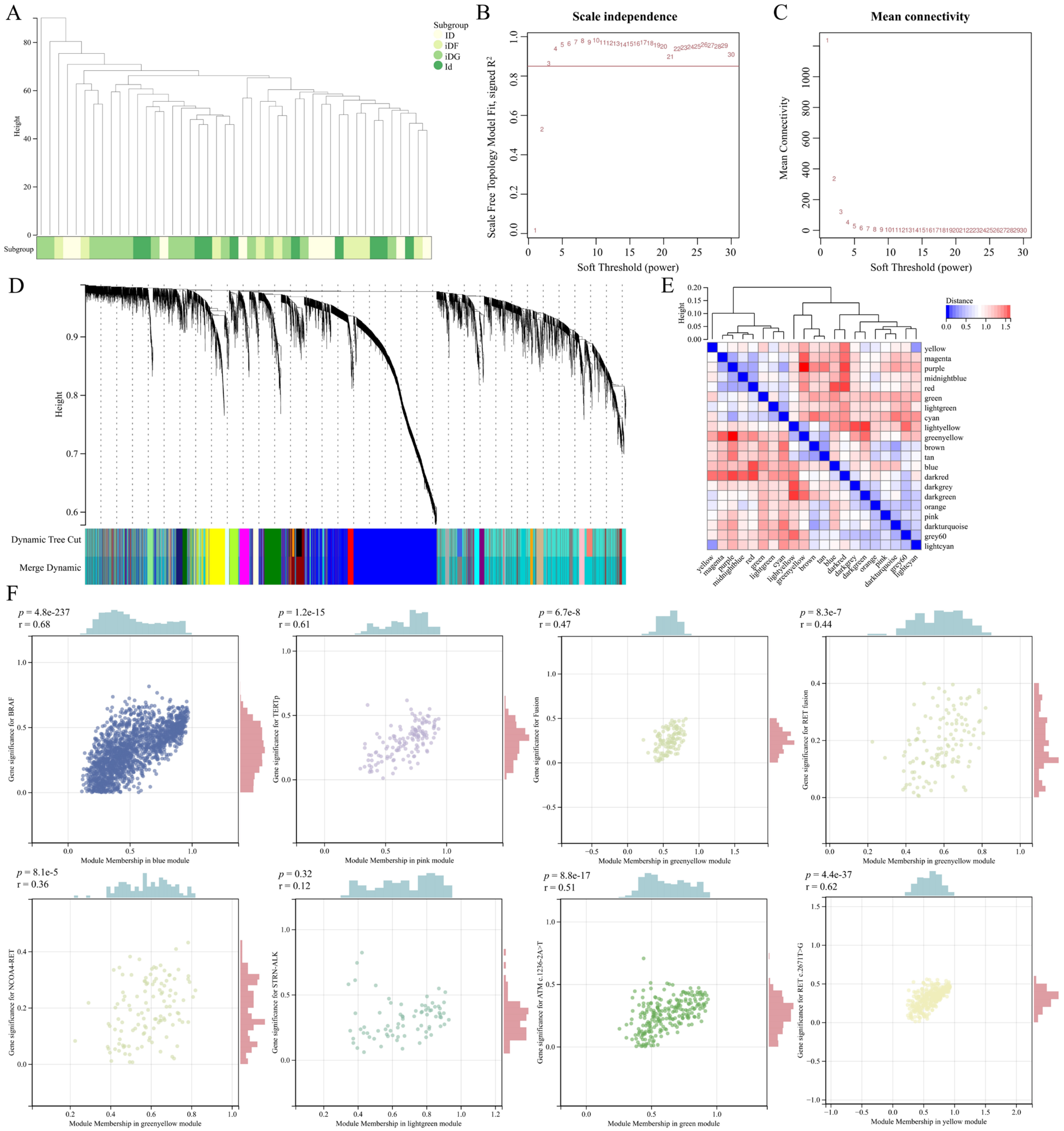

Supplementary figure 10. Weighted gene co-expression network analysis. (A) Unsupervised hierarchical clustering of samples with the Euclidean distance and average clustering methods. (B) Scale independence analysis. (C) Mean connectivity analysis. When the power was 3, the scale-free $R^2$ was > 0.85 for the first time, and the mean connectivity curve gradually stabilized. The soft thresholding power was thus set to 3. (D) Clustering dendrogram of all genes. (E) Module eigengene adjacency heatmap. The gray module was the collection of genes that were not classified into any other modules. (F) Scatter plots of gene variances and modules.

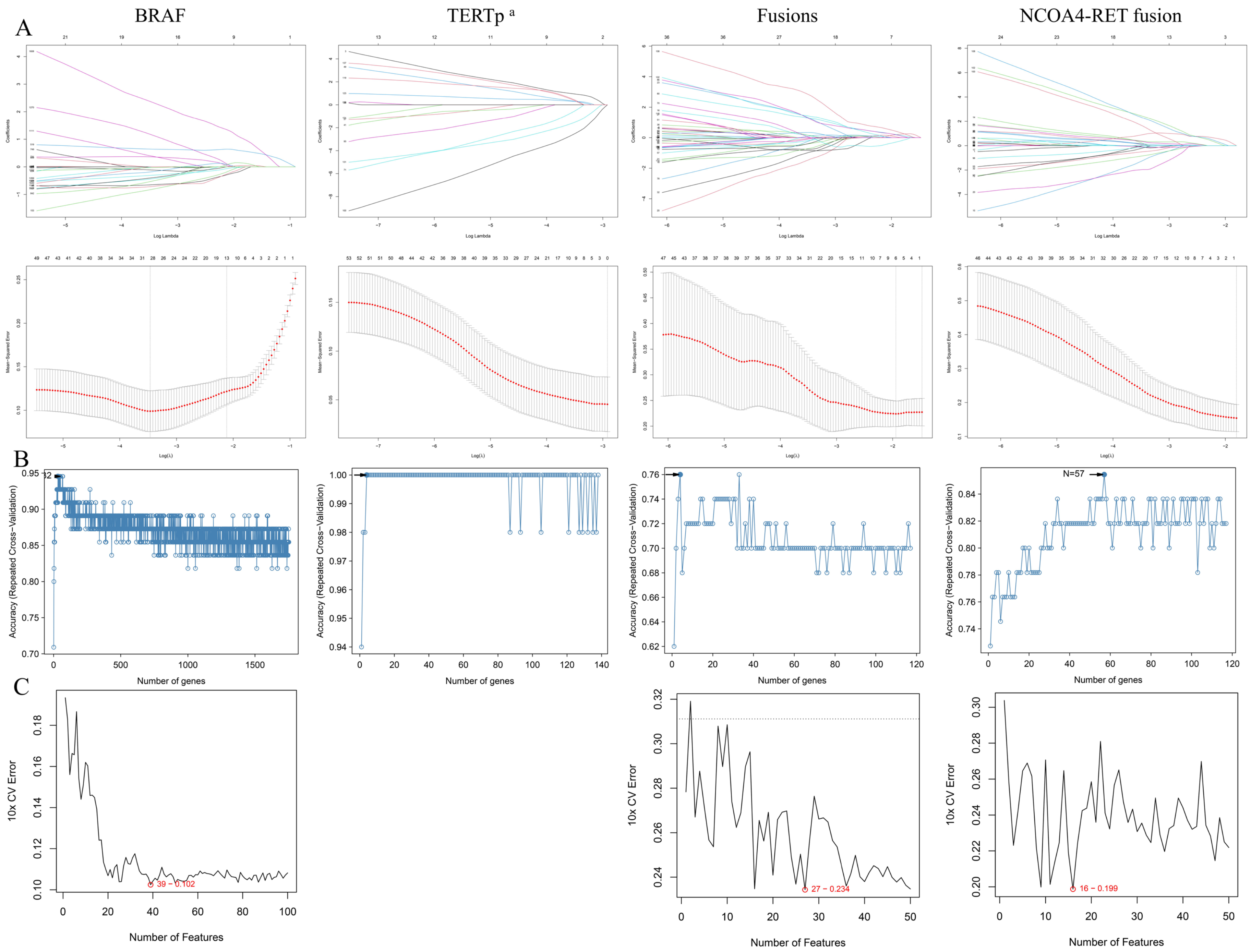
A
BRAF
TERTp a
Fusions
NCOA4-RET fusion
Log Lambda
Coefficients
Mean−Squared Error
Log(λ)
B
Accuracy (Repeated Cross−Validation)
Number of genes
N=57
C
10x CV Error
Number of Features
39 − 0.102
27 − 0.234
16 − 0.199

Supplementary figure 11. Identification of biomarker proteins using machine learning. (A) The reduction in variables and adjustment of the β value during LASSO regression. (B) The accuracy plots along with the numbers of genes in the RF models. (C) The error plots along with the numbers of genes in the SVM models. The four columns represent *BRAF*, *TERTp*, gene fusions and *NCOA4-RET* fusion from left to right.

[a] The low number of *TERTp*-positive samples (n = 6) prevented model construction and error analysis for *TERTp* mutations via the SVM algorithm.

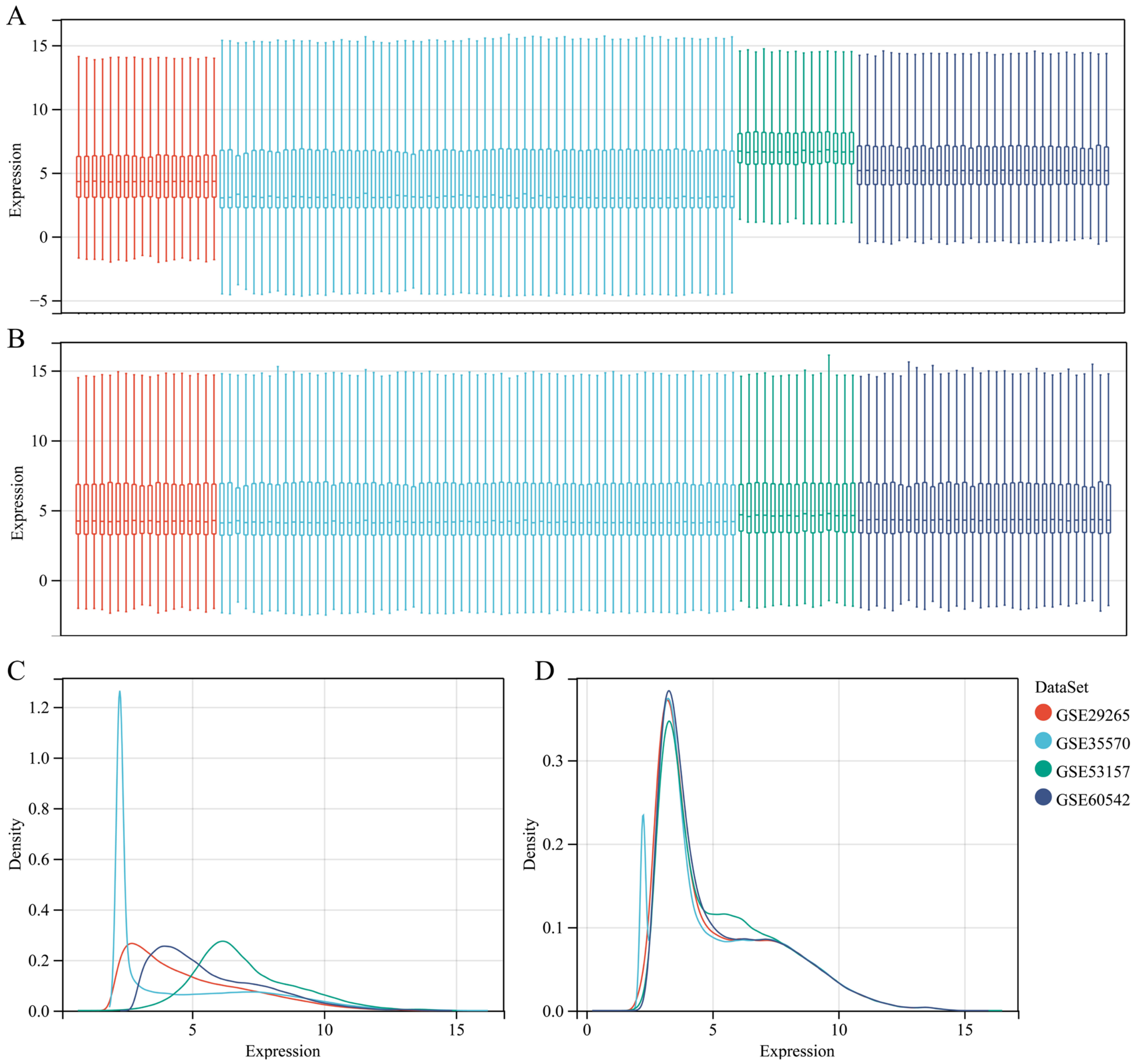

Supplementary figure 12. Batch effect removal of the four external datasets. Boxplots of the expression of the samples before (A) and after (B) batch effect removal. Density plots of expression within each dataset before (C) and after (D) batch effect removal.

15. Sun Y, Li L, Zhou Y, Ge W, Wang H, Wu R, et al. Stratification of follicular thyroid tumours using data-independent acquisition proteomics and a comprehensive thyroid tissue spectral library. Molecular

oncology. 2022;16(8):1611-24.
16. Cai X, Xue Z, Wu C, Sun R, Qian L, Yue L, et al. High-throughput proteomic sample preparation using pressure cycling technology. Nature Protocols. 2022;17(10):2307-25.
17. Demichev V, Messner CB, Vernardis SI, Lilley KS, Ralser M. DIA-NN: neural networks and interference correction enable deep proteome coverage in high throughput. Nature Methods. 2020;17(1):41-+.
18. Bateman A, Martin M-J, Orchard S, Magrane M, Ahmad S, Alpi E, et al. UniProt: the Universal Protein Knowledgebase in 2023. Nucleic Acids Research. 2022;51(D1):D523-D31.
19. Xiong Y, Li X, Li L, Li D, Yan L, Li X, et al. Application of biomarkers in the diagnosis of uncertain samples of core needle biopsy of thyroid nodules. Virchows Archiv. 2021;479(5):961-74.
20. Landrum MJ, Lee JM, Benson M, Brown G, Chao C, Chitipiralla S, et al. ClinVar: public archive of interpretations of clinically relevant variants. Nucleic Acids Research. 2016;44(D1):D862-D8.
21. Tate JG, Bamford S, Jubb HC, Sondka Z, Beare DM, Bindal N, et al. COSMIC: the Catalogue Of Somatic Mutations In Cancer. Nucleic Acids Research. 2019;47(D1):D941-D7.
22. Subramanian A, Tamayo P, Mootha VK, Mukherjee S, Ebert BL, Gillette MA, et al. Gene set enrichment analysis: A knowledge-based approach for interpreting genome-wide expression profiles. Proceedings of the National Academy of Sciences of the United States of America. 2005;102(43):15545-50.
23. Liberzon A, Subramanian A, Pinchback R, Thorvaldsdottir H, Tamayo P, Mesirov JP. Molecular signatures database (MSigDB) 3.0. Bioinformatics. 2011;27(12):1739-40.
24. Haenzelmann S, Castelo R, Guinney J. GSVA: gene set variation analysis for microarray and RNA-Seq data. Bmc Bioinformatics. 2013;14:7.
25. Zhou Y, Zhou B, Pache L, Chang M, Khodabakhshi AH, Tanaseichuk O, et al. Metascape provides a biologist-oriented resource for the analysis of systems-level datasets. Nature Communications. 2019;10:1523.
26. Kraemer A, Green J, Pollard J, Jr., Tugendreich S. Causal analysis approaches in Ingenuity Pathway Analysis. Bioinformatics. 2014;30(4):523-30.
27. Newman AM, Steen CB, Liu CL, Gentles AJ, Chaudhuri AA, Scherer F, et al. Determining cell type abundance and expression from bulk tissues with digital cytometry. Nature Biotechnology. 2019;37(7):773-+.
28. Yoshihara K, Shahmoradgoli M, Martinez E, Vegesna R, Kim H, Torres-Garcia W, et al. Inferring tumour purity and stromal and immune cell admixture from expression data. Nature Communications. 2013;4:2612.
29. Rooney MS, Shukla SA, Wu CJ, Getz G, Hacohen N. Molecular and Genetic Properties of Tumors Associated with Local Immune Cytolytic Activity. Cell. 2015;160(1-2):48-61.
30. Barbie DA, Tamayo P, Boehm JS, Kim SY, Moody SE, Dunn IF, et al. Systematic RNA interference reveals that oncogenic KRAS-driven cancers require TBK1. Nature. 2009;462(7269):108-U22.
31. Charoentong P, Finotello F, Angelova M, Mayer C, Efremova M, Rieder D, et al. Pan-cancer Immunogenomic Analyses Reveal Genotype-Immunophenotype Relationships and Predictors of Response to Checkpoint Blockade. Cell Reports. 2017;18(1):248-62.
32. Senbabaoglu Y, Gejman RS, Winer AG, Liu M, Van Allen EM, de Velasco G, et al. Tumor immune microenvironment characterization in clear cell renal cell carcinoma identifies prognostic and immunotherapeutically relevant messenger RNA signatures. Genome Biology. 2016;17:231.

33. Agrawal N, Akbani R, Aksoy BA, Ally A, Arachchi H, Asa SL, et al. Integrated Genomic Characterization of Papillary Thyroid Carcinoma. Cell. 2014;159(3):676-90.
34. Pratilas CA, Taylor BS, Ye Q, Viale A, Sander C, Solit DB, et al. (V600E)BRAF is associated with disabled feedback inhibition of RAF-MEK signaling and elevated transcriptional output of the pathway. Proceedings of the National Academy of Sciences of the United States of America. 2009;106(11):4519-24.
35. Langfelder P, Horvath S. WGCNA: an R package for weighted correlation network analysis. Bmc Bioinformatics. 2008;9:559.
36. Breiman L. Random forests. Machine Learning. 2001;45(1):5-32.
37. Engebretsen S, Bohlin J. Statistical predictions with glmnet. Clinical Epigenetics. 2019;11(1):123.
38. Sanz H, Valim C, Vegas E, Oller JM, Reverter F. SVM-RFE: selection and visualization of the most relevant features through non-linear kernels. Bmc Bioinformatics. 2018;19:432.
39. Colombo C, Minna E, Gargiuli C, Muzza M, Dugo M, De Cecco L, et al. The molecular and gene/miRNA expression profiles of radioiodine resistant papillary thyroid cancer. Journal of Experimental & Clinical Cancer Research. 2020;39(1):245.
40. Barrett T, Wilhite SE, Ledoux P, Evangelista C, Kim IF, Tomashevsky M, et al. NCBI GEO: archive for functional genomics data sets-update. Nucleic Acids Research. 2013;41(D1):D991-D5.
41. Jang YE, Jang I, Kim S, Cho S, Kim D, Kim K, et al. ChimerDB 4.0: an updated and expanded database of fusion genes. Nucleic Acids Research. 2020;48(D1):D817-D24.
42. Handkiewicz-Junak D, Swierniak M, Rusinek D, Oczko-Wojciechowska M, Dom G, Maenhaut C, et al. Gene signature of the post-Chernobyl papillary thyroid cancer. European Journal of Nuclear Medicine and Molecular Imaging. 2016;43(7):1267-77.
43. Pita JM, Banito A, Cavaco BM, Leite V. Gene expression profiling associated with the progression to poorly differentiated thyroid carcinomas. British Journal of Cancer. 2009;101(10):1782-91.
44. Tarabichi M, Saiselet M, Tresallet C, Hoang C, Larsimont D, Andry G, et al. Revisiting the transcriptional analysis of primary tumours and associated nodal metastases with enhanced biological and statistical controls: application to thyroid cancer. British Journal of Cancer. 2015;112(10):1665-74.
45. Tang Z, Kang B, Li C, Chen T, Zhang Z. GEPIA2: an enhanced web server for large-scale expression profiling and interactive analysis. Nucleic Acids Research. 2019;47(W1):W556-W60.
46. Davis MDP, Perniciaro C, Dahl PR, Randle HW, McEvoy MT, Leiferman KM. Exaggerated arthropod-bite lesions in patients with chronic lymphocytic leukemia: A clinical, histopathologic, and immunopathologic study of eight patients. Journal of the American Academy of Dermatology. 1998;39(1):27-35.
47. Gonzalez-Juarrero M, Orme IM. Characterization of murine lung dendritic cells infected with Mycobacterium tuberculosis. Infection and Immunity. 2001;69(2):1127-33.
48. Koethe S, Zander L, Koster S, Annan A, Ebenfelt A, Spencer J, et al. Pivotal Advance: CD45RB glycosylation is specifically regulated during human peripheral B cell differentiation. Journal of Leukocyte Biology. 2011;90(1):5-19.
49. Li S, Xu F, Zhang J, Wang L, Zheng Y, Wu X, et al. Tumor-associated macrophages remodeling EMT and predicting survival in colorectal carcinoma. Oncoimmunology. 2018;7(2):e1380765.
